# Adult current and former nicotine users’ and adolescents’ views on vape-free places: findings from UK cross-sectional surveys

**DOI:** 10.64898/2026.07.06.26357368

**Authors:** Allison Ford, Catherine Best, Crawford Moodie, Georgia Alexandrou, Anne Marie MacKintosh

## Abstract

**Introduction:** The Tobacco and Vapes Act 2026 provides the UK government powers to ban vaping in public places. Proposals include extending existing indoor smoke-free legislation to also being vape-free and making public children’s playgrounds and outdoor areas of education settings vape-free. We examined adults’ and adolescents’ views on vape-free places.

**Methods:** Two UK-wide cross-sectional online surveys were conducted in 2026, one with adult (18+) current and former nicotine users (n=2,851), and one with adolescents aged 11-17 years (n=2,123). We measured (1) perceived acceptability of vaping in public places, (2) views on whether vapes should/should not be allowed in public places, and (3) perceived likelihood of public compliance with vaping bans.

**Results:** Adults considered it unacceptable to vape on public transport (85.9%), on school grounds (outdoors) (83.6%), outside hospital entrances (59.2%), and inside pubs (57.8%) and nightclubs (52.5%). A minority considered it unacceptable to vape at open air playparks (40.6%). Most adolescents viewed vaping as unacceptable in all locations. Perceived acceptability of vaping in each place was associated with current vaping and/or smoking. Adults and adolescents believed vaping should not be allowed on public transport (88.9%; 93.8%) or outdoors on school grounds (85.8%; 93.5%). Adults and adolescents perceived likely compliance on public transport, school grounds and in pubs, but not in nightclubs, outside hospital entrances and at open air playparks.

**Conclusion:** The findings indicate public support for some vape-free places among adults and adolescents, particularly on public transport and school grounds, and less so for a ban at open air playparks.

**IMPLICATIONS:** This study focused on the views of current and former nicotine users and adolescents across the UK. Those most affected by vape-free policies may benefit from greater understanding of the rationale behind proposals and convincing of the practical feasibility of banning vaping in some places.

## INTRODUCTION

The United Kingdom (UK) Government’s Tobacco and Vapes Act 2026 is intended to reduce preventable deaths from smoking and help tackle youth vaping.^1,2^ Measures including the “smokefree generation”, which prohibits the sale of tobacco to anyone born on or after 1st January 2009, and a ban on the advertising and sponsorship of vapes and other nicotine products, will be enacted on the face of the bill and implemented in 2027.^3^ The Act provides the government powers to introduce further regulatory measures on vaping and nicotine products such as restricting flavours, packaging and retail display, and a retailer licensing scheme.^1,3^

Within this legislative framework, another regulatory measure was proposed to ban vaping in public places. This was included within the consultation on ‘Smoke-free, heated tobacco-free and vape-free places in England’, which ran from February-May 2026; separate consultations will be conducted in Scotland, Wales, and Northern Ireland. It included proposals to (i) extend existing bans on smoking indoors to some outdoor areas, (ii) make existing indoor smoke-free places heated tobacco- and vape-free, (iii) make all outdoor areas becoming smoke-free also heated tobacco-free, and (iv) make public children’s playgrounds and the outdoor areas of education settings vape-free.^4^ The indoor and outdoor vape-free proposals were presented in the context of World Health Organization advice that second-hand vapour poses potential health risks to people who vape and those around them from raised concentrations of particulate matter which contain nicotine and other potentially toxic substances.^5,6^ The proposals were also framed around the objective to reduce the appeal and uptake of vaping among young people and non-smoking adults through minimising the visibility, and thereby norms, of vaping among these groups, while being mindful of the role vapes play in smoking cessation for adult smokers.^4,6,7^

Unlike for smoke-free places, where there is a large international evidence base examining public support, compliance, and impacts (e.g. reduced exposure to harmful second-hand smoke, improved cardiovascular health),^8-11^ there is limited evidence on potential impacts of vape-free places. Studies exploring indoor second-hand exposure to e-cigarette vapour have found associations with self-reported respiratory problems such as bronchitis symptoms, shortness of breath and asthma exacerbation in young people.^12,13^ Studies examining biomarkers of second-hand toxicant exposure show higher nicotine absorption among children and adults exposed to vapour compared with those not exposed,^14-16^ although at levels lower than those exposed to second-hand smoke.^14^ While evidence also suggests a higher level of cobalt (out of 27 biomarkers of metals),^16^ and the inflammatory cytokine IL-1β,^15^ in those exposed versus not exposed, the overall harm from second-hand vapour exposure is likely far lower than harm from second-hand smoking.^17^

There may be impacts from seeing others vape. A systematic review concluded that social interactions and social norms around vaping, especially among peers, friends and family, play a role in vaping decisions.^18^ Seeing anyone vape was found to be a predictor of vaping initiation among youth and young adults.^19^ A study examining household vaping bans found that in households where parents had introduced a ban, there were lower odds of young people vaping.^20^ Social influence and observational learning may therefore play a role in vaping uptake, similar to how tobacco social norms have played a role in youth smoking.^18^

It is critical to understand public views on potential policies. Public support can influence policymakers in decisions to enact policies.^21^ Public acceptability of a policy can affect implementation and effectiveness.^22,23^ Assessing public views can identify where efforts are required to increase policy acceptability,^24^ given the important role of acceptability in compliance.^25^ While there is strong public support across Great Britain for extending smoke-free legislation to additional places,^26^ far less is known about support for vape-free places. Most studies are with adults from the United States (US) and Australia. In a survey of US adults just under half believed vaping should ‘never be allowed’ in restaurants, a third that it should be banned entirely indoors at bars, casinos and clubs, while only a quarter supported a ban in parks.^27^ Five years later, public support in the US had risen for banning vaping in all indoor public places (82.9%), restaurants (86.5%), and bars (76.1%).^28^ Support was similarly high among Australian adults, with most (83%) agreeing that vaping should not be allowed on public transport, in pubs, restaurants, or other indoor venues.^29^ Studies examining support among adults who currently smoke,^27,28,30^, currently vape,^28^ or have ever vaped,^27^ consistently report lower levels of support for vape-free policies compared with non-users. In one study 41.2% of people reporting current smoking supported prohibiting indoor vaping.^30^ In another study, more than half of people reporting current vaping supported prohibiting vaping in restaurants.^28^

To our knowledge no study has explored the UK population’s views on indoor and outdoor vape-free places. We present findings from two surveys exploring adult current and former nicotine users’ and adolescents’ views on vape-free places. Specifically, we explore (1) perceived acceptability of vaping in public places, (2) views on whether vapes should or should not be allowed in public places, and (3) perceived likelihood of public compliance with bans on vaping in public places.

## METHODS

### Design

Data were collected in two UK-wide cross-sectional online surveys, one with adult (18+) current and former nicotine users, and one with adolescents (11-17 years). We focused on adults who currently or have previously used nicotine products given the importance of understanding how public opinions vary across behavioural domains,^22^ particularly among those most likely affected by vape-free policy. We included people who currently or have previously smoked as a relevant stakeholder group given vaping’s role in smoking cessation.^7,30^ We included adolescents because of the popularity of vapes among this group.^2^ YouGov, a market research company, was commissioned to host the survey and recruit participants. Ethical approval was granted by the University of Stirling General University Ethics Panel (GUEP 2025 23794 18341).

### Sample and recruitment

YouGov recruited participants from their UK online panels. Adults were targeted according to data held on their smoking/vaping status and invited by email to participate in the study. For 11–15-year-olds, YouGov targeted panel members known to have a child of relevant age and invited the child via their parent. Participants aged 16-17 years were approached directly if an existing panel member, or via their parent.

### Data collection

Data were collected between April and May 2026 for adults and April and June 2026 for adolescents. Informed consent was required prior to survey completion, including parental/guardian consent for 11–15-year-olds.

### Development of measures to explore perceptions of vape-free places

Measures around vape-free places were developed through six in-person public involvement discussion groups, four with adults, one with older adolescents (16-17 years) and one with younger adolescents (14-15 years). This enabled us to draft questions according to how members of the public viewed the issue of vape-free. Drafts of questions and response options were tested and refined in 16 individual cognitive interviews with adults and adolescents to ensure relevance, comprehension, and suitability. Questions were designed to reduce bias by using balanced questioning and response options. The development phase of survey items was conducted October-November 2025.

### Measures

#### Demographics

Age, gender, social grade and country were obtained from information that YouGov hold about panel members. Social grade was determined by the occupation of the chief income earner in the household (ABC1=middle class, C2DE=working class).

#### Smoking and vaping status

Adult and adolescents’ vaping status was determined through asking ‘*Which of these best describes whether or not you have ever used or tried vapes? (1) I have never used vapes, (2) I have only ever tried vapes once or twice, (3) I have used vapes in the past but I never use them now, (4) I occasionally use vapes (less than once a month), (5) I use vapes at least once a month, (6) I use vapes at least once a week, (7) I use vapes every day’*. Those responding 4-7 were categorised as currently vaping.

Adult smoking status was determined through asking ‘*Which of the following best describes you? (1) I have never smoked, not even tried a puff or two, (2) I tried smoking in the past but I never smoke cigarettes (including hand-rolled) now, (3) I used to smoke but I never smoke cigarettes (including hand-rolled) now, (4) I smoke cigarettes (including hand-rolled) but not every day, (5) I smoke cigarettes (including hand-rolled) every day’* Those responding 4-5 were categorised as currently smoking.

A four-category classification of adults’ vaping/smoking status was established from the vaping and smoking questions: 1) Exclusively vaping, 2) Exclusively smoking, 3) Dual use, and 4) Not currently vaping/smoking.

#### Perceived acceptability of vaping in public places

All participants were asked ‘*On a scale of one to five where 1 is very acceptable and 5 is very unacceptable, how acceptable or unacceptable do you think it is for people to vape in the following places? (1) Open air playparks, (2) On public transport, (3) At hospital entrances (outdoors), (4) Inside nightclubs, (5) Pubs (indoors), (6) On school grounds (outdoors)’* Responses were given on a 1-5 rating scale from (1) Very acceptable to (5) Very unacceptable, with a *‘Not sure’* response category.

#### Views on banning vaping in public places

All participants were asked ‘*Do you think that people should or should not be allowed to vape in the following places? (1) Open air playparks, (2) On public transport, (3) At hospital entrances (outdoors), (4) Inside nightclubs, (5) Pubs (indoors), (6) On school grounds (outdoors)’* Response options were ‘*Should be allowed*’, ‘*Should not be allowed*’, and ‘*Not sure’*.

#### Perceived compliance with bans on vaping in public places

All participants were asked ‘*If vaping was banned in the following places do you think people would be likely or unlikely to obey the rules? (1) Open air playparks, (2) On public transport, (3) At hospital entrances (outdoors), (4) Inside nightclubs, (5) Pubs (indoors), (6) On school grounds (outdoors)*. Response options were ‘*Very likely to obey’*, ‘*Quite likely to obey*’, *Quite unlikely to obey’, Very unlikely to obey’*, and ‘*Not Sure’*.

### Analysis

Data was analysed using SPSSv31. For adolescents, descriptive statistics were weighted to be representative of the UK population of 11–17-year-olds. YouGov generated weights using Random Iterative Method weighting to bring the demographic profile in line with the population in terms of age crossed by gender and region. No weights were generated for adults, as a sample with current and past smoking and/or vaping experience was sought rather than a general population sample.

Logistic regression analyses were undertaken to explore the characteristics of people who might be resistant to restrictions on vaping in each place. In both the adult and adolescent samples logistic regressions, controlling for vaping behaviour, age, gender (female vs. male), social grade (C2DE vs. ABC1) and country (each nation vs. England), were run to examine differences in likelihood of each outcome for each place. The adult and adolescent logistic regression analyses differed in terms of the behaviour variable and age variable included. For adults, age was available as a categorical variable and each age category was compared against the combined ages below it (e.g. 25-34 years vs. 18-24 years and 45-54 years vs. 18-44 years). We used a four-category variable of current vaping/smoking, with the reference category being the group who did not currently vape or smoke. Each remaining category (dual use, exclusively vaping, exclusively smoking) was compared against the non-user group. For adolescents, actual age was included as an integer value, subject to meeting the required linearity of the logit conditions. Due to the smaller sample and fewer people who vaped and/or smoked, logistic regressions examined the association between each outcome variable and vaping status. The reference category was those who did not currently vape. The binary outcome variables, for each of the six places, were: whether vaping was perceived to be acceptable (vs. neutral, acceptable or not sure); whether people thought vaping should be allowed (vs. not allowed or not sure); and whether they thought it was unlikely that people would obey vaping restrictions (vs. likely to obey or not sure). All logistic regression results are presented in Supplementary Tables 1a-6b. Within the text we draw out differences significant at p<0.01.

## RESULTS

A total of 2,851 adults and 2,123 adolescents were included in the study. Respondent characteristics are presented in Table 1.

**Table 1:**
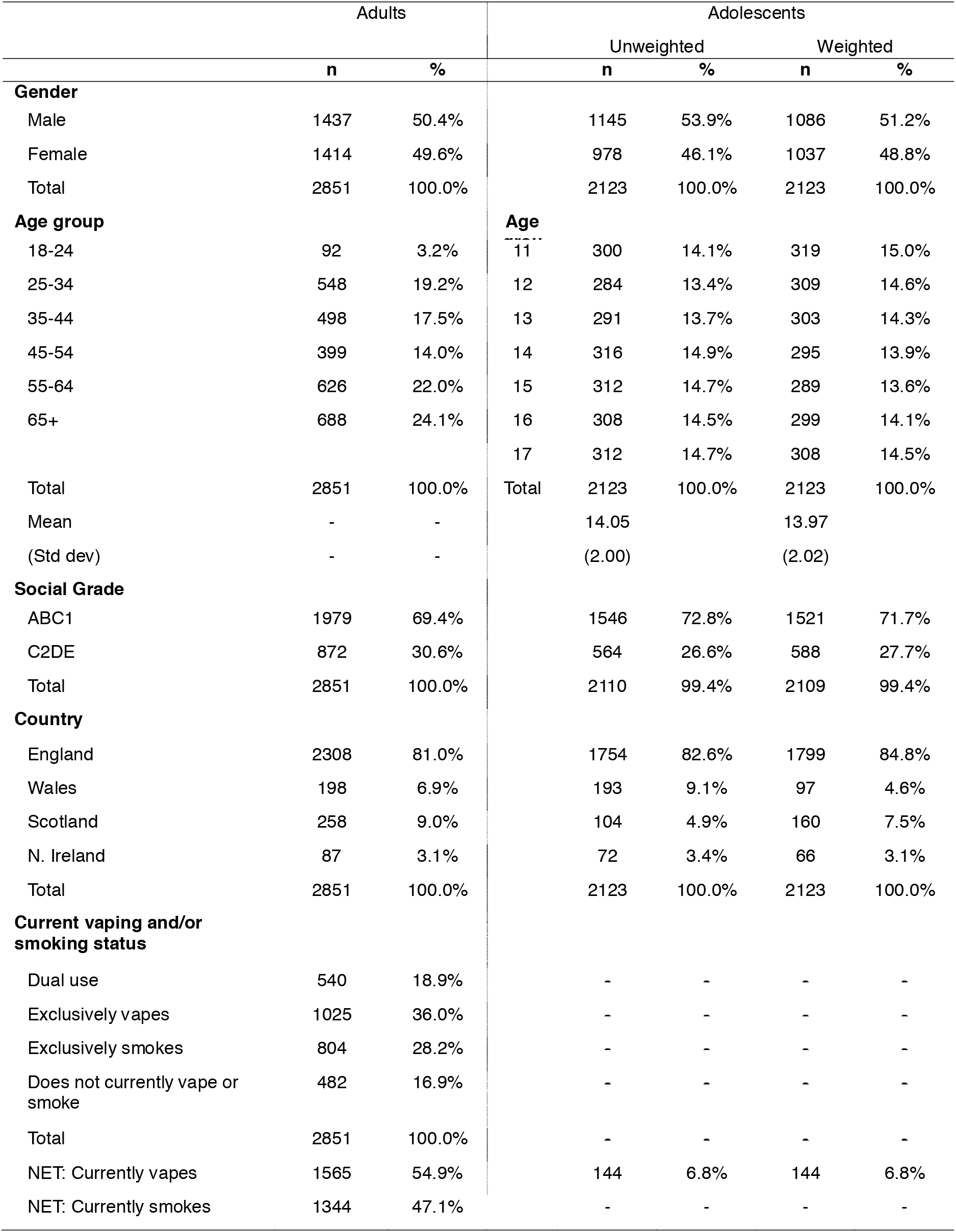
Sample characteristics: Adults (Current and former nicotine users) and Adolescents aged 11 to 17 years.

### Perceived acceptability of vaping in different places

Adult current and former nicotine users perceived it to be unacceptable to vape in five of the six places they were asked about (Figure 1a; Supplementary Table 1a), particularly on public transport (85.9%) and school grounds (83.6%) and to a lesser extent at hospital entrances (59.2%) or in pubs (57.8%) and nightclubs (52.5%). Similar proportions viewed vaping in open air playparks to be either unacceptable (40.6%) or acceptable (37.3%).

**Figure 1:**
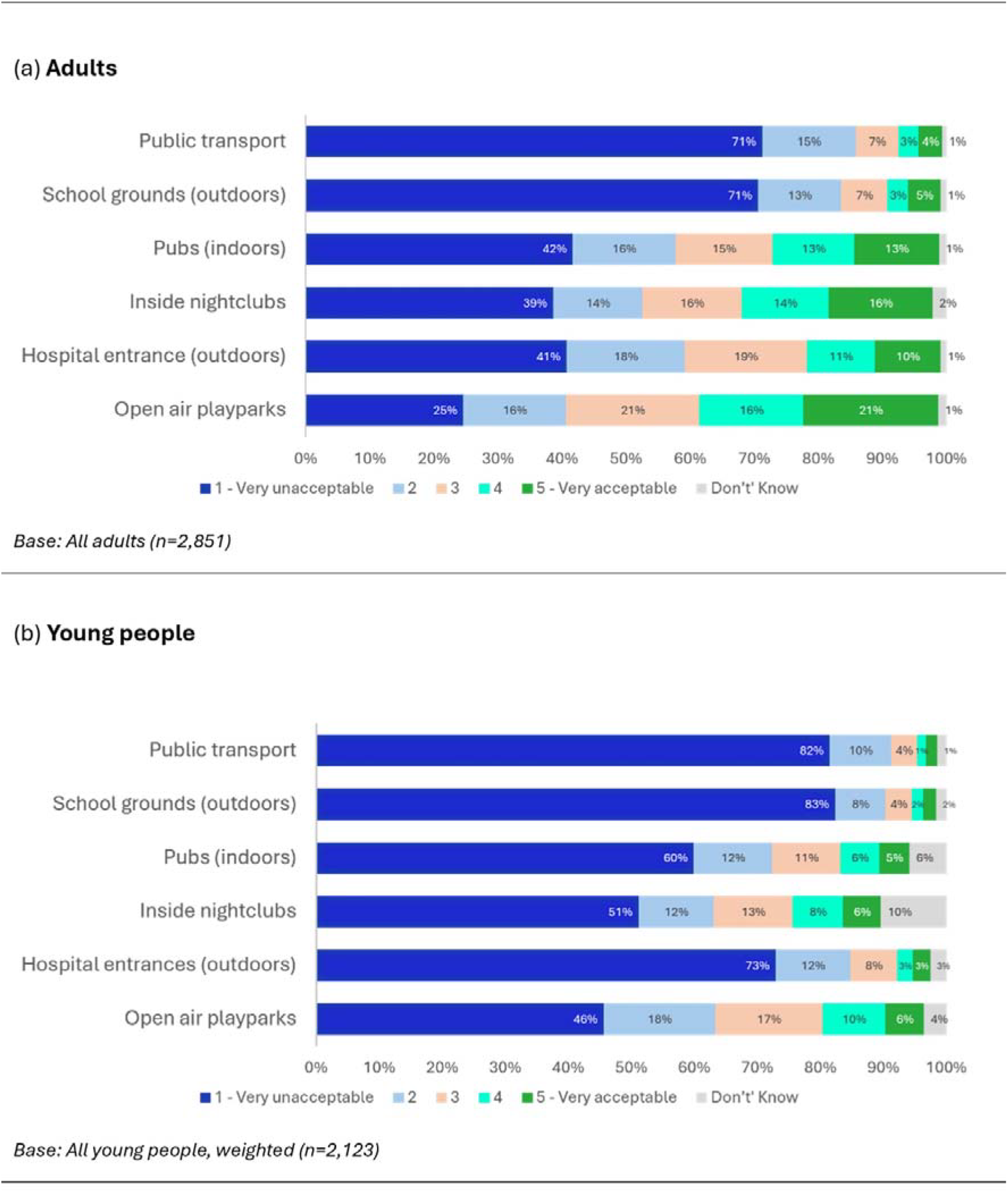
Perceptions of the acceptability of vaping in different locations.

Most adolescents perceived vaping to be unacceptable in all six places (Figure 1b; Supplementary Table 1b), particularly on public transport (91.2%), school grounds (90.5%) and at hospital entrances (84.7%) and to a lesser extent in pubs (72.5%), nightclubs (63.2%) and open air playparks (63.4%).

### Views on whether vaping should or should not be allowed in different places

For five of the six places, adults thought that vaping should not be allowed (Figure 2a; Supplementary Table 2a), particularly on public transport (88.9%) or on school grounds (85.8%). Most also thought that it should not be allowed in pubs (59.9%), at hospital entrances (58.6%) or in nightclubs (55.0%). For open air playparks, more adults thought that vaping should be allowed (49.4%) than not allowed (41.3%).

**Figure 2:**
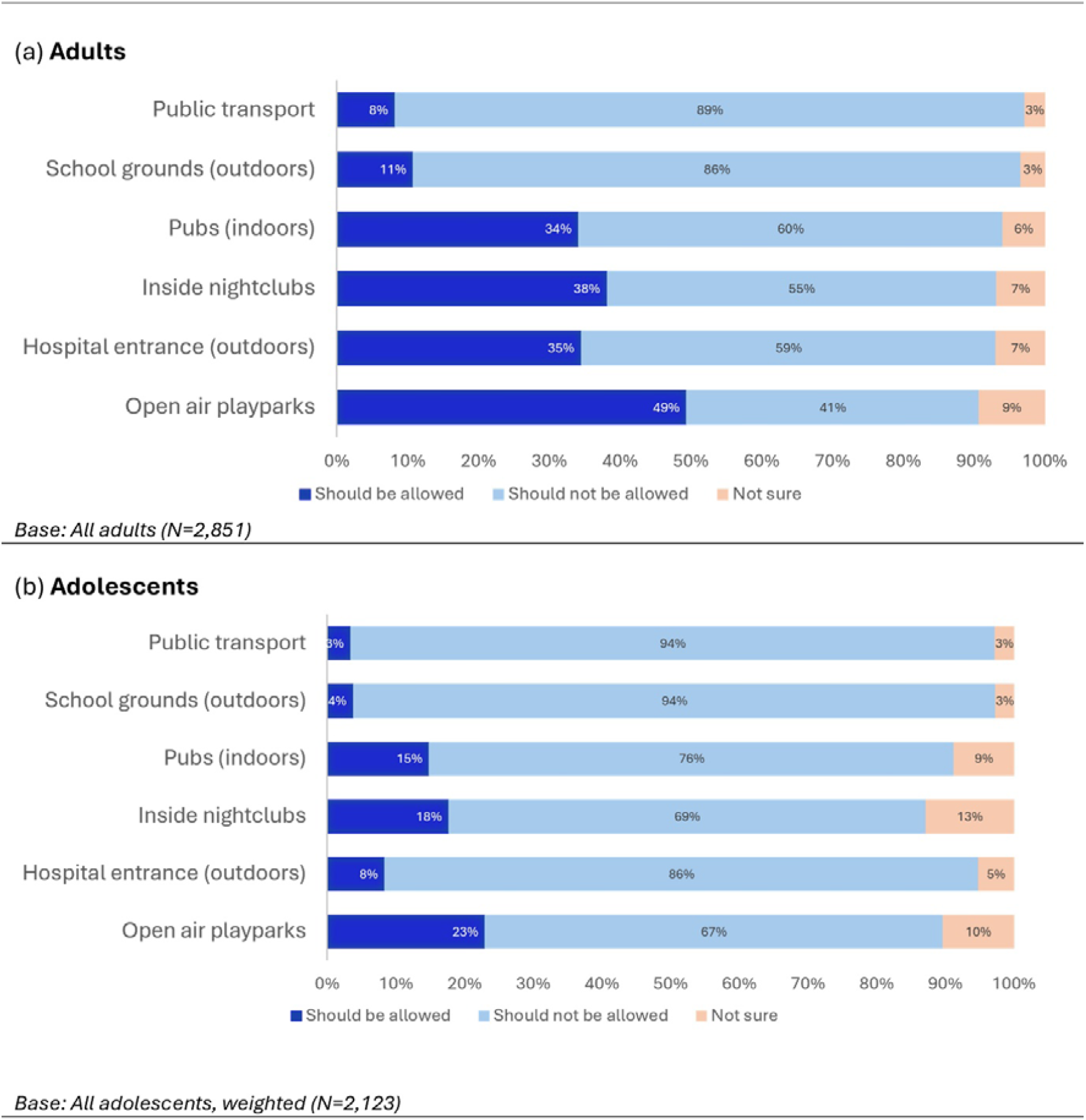
Views on whether vaping should or should not be allowed in different places.

Most adolescents thought that vaping should not be allowed in any of the six places (Figure 2b; Supplementary Table 2b). However, even among adolescents, there were mixed views with more than a fifth thinking it should be allowed in open air playparks (23.0%). Similarly, at least one in seven thought it should be allowed in pubs (14.9%) and in nightclubs (17.7%).

#### Differences by behaviour and demographics - adults

Logistic regression analyses indicated that, for each of the six locations, adults who currently vaped and/or smoked (whether dual using, exclusively vaping or exclusively smoking) were more likely than non-current users to perceive it as acceptable to vape in each place (Supplementary Table 3a).

For each of the six places, females were less likely than males to think it acceptable to vape. Only for nightclubs and pubs were age differences evident. For nightclubs, the likelihood of considering it acceptable to vape decreased with increasing age category. For example, 35– 44-year-olds were less likely than 18–34-year-olds to consider it acceptable (Adjusted Odds Ratio (AOR) 0.55, 95% CI [0.41-0.74], p<0.001). Those aged 65 and over were less likely than 18–64-year-olds to consider it acceptable to vape in pubs (AOR 0.66, 95% CI [0.52-0.84], p<0.001).

Logistic regression with the outcome variable being ‘vaping should be allowed’ in each location showed the same pattern of differences in terms of behaviour, gender, and age (Supplementary Table 4a).

#### Differences by behaviour and demographics – adolescents

Similar to adults, adolescents who currently vaped were more likely than those not vaping to perceive it acceptable to vape in each place (Supplementary Tables 3b). Females were less likely than males to perceive it acceptable to vape in nightclubs (AOR 0.66, 95% CI [0.50-0.86], p<0.01) and pubs (AOR 0.62, 95% CI [0.46-0.83], p<0.01). Logistic regression with the outcome variable being ‘vaping should be allowed’ in each location showed the same pattern of differences in terms of behaviour and gender (Supplementary Table 4b)

### Views on likely compliance with vaping restrictions in different places

Most adults felt that people would obey the rules if vaping restrictions were introduced on public transport (72.6%), on school grounds (63.4%) and in pubs (57.7%). Fewer thought that people would obey vaping restrictions in nightclubs (42.2%), at hospital entrances (35.5%) and at open air playparks (25.7%). (Figure 3a; Supplementary Table 5a).

**Figure 3:**
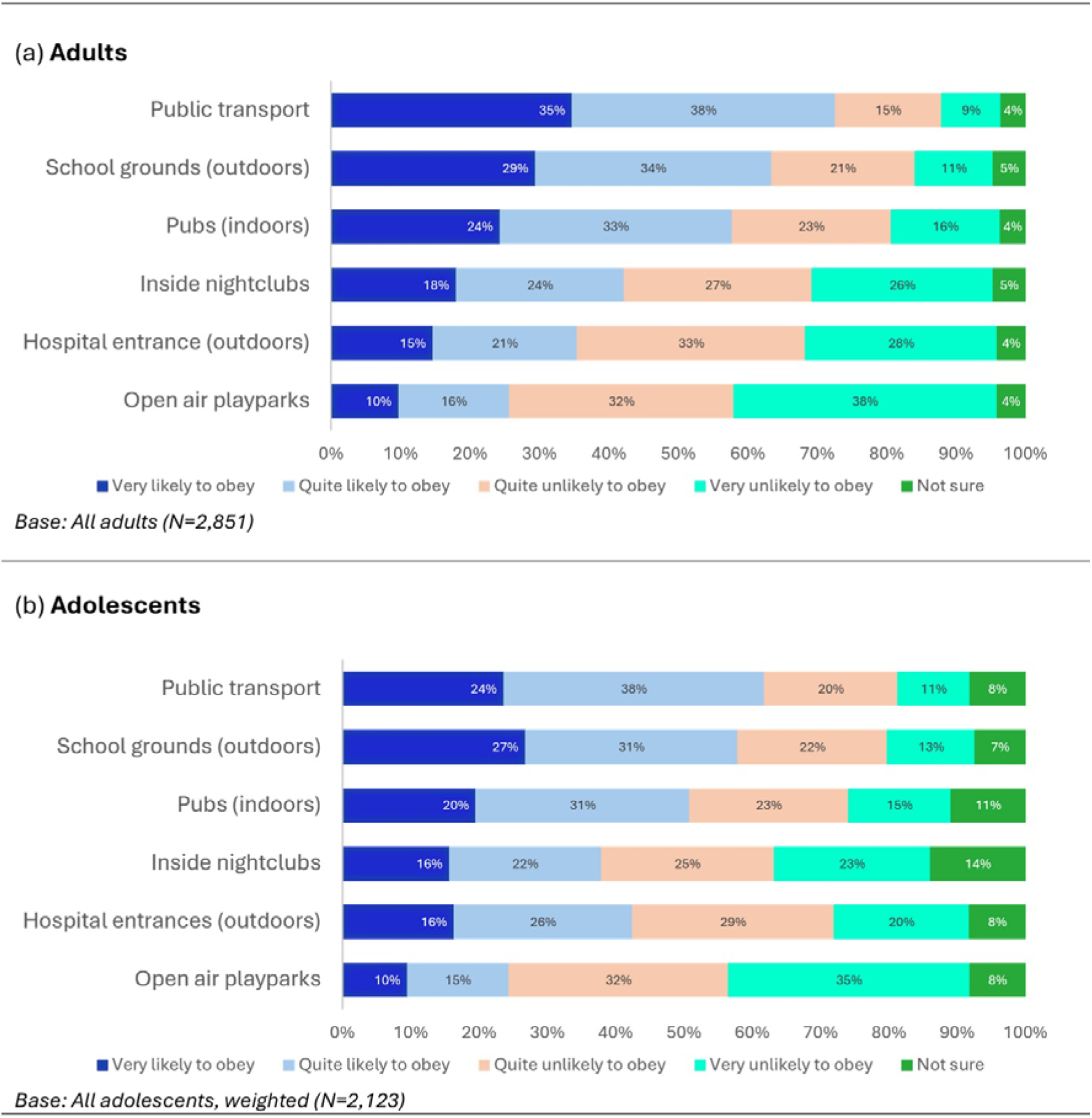
Perceptions of likelihood of compliance if vaping was banned in the following places.

Most adolescents also felt that people would obey the rules on public transport (61.8%), on school grounds (58.0%) and in pubs (50.9%), with fewer thinking that people would obey vaping restrictions in nightclubs (37.9%), at hospital entrances (42.5%) and at open air playparks (24.4%). (Figure 3b; Supplementary Table 5b).

#### Differences by behaviour and demographics – adults

Logistic regression analyses indicated some variation by adults’ current vaping and/or smoking status (Supplementary Table 6a). Adults who currently vaped and/or smoked (i.e. those dual using; exclusively vaping; exclusively smoking) were less likely than non-current users to doubt compliance at open air playparks and school grounds. For hospital entrances, dual users (AOR 0.69, 95% CI [0.53-0.89], p<0.01) were less likely than non-users to doubt compliance. Dual users and those exclusively vaping were less likely to doubt compliance on public transport.

Compared with males, females were more likely to doubt compliance at hospital entrances (AOR 1.25, 95% CI [1.07-1.45], p<0.01) and less likely to doubt compliance on public transport (AOR 0.79, 95% CI [0.66-0.94], p<0.01). Differences by age were observed for all places except school grounds and playparks. These tended to show that older participants were less likely to doubt compliance. The exception being hospital entrances where older participants were more likely to doubt compliance. Adults from social grade C2DE (working class) were more likely than those from ABC1 (middle class) to doubt compliance at hospital entrances (AOR 1.39, 95% CI [1.17-1.66], p<0.001) and on school grounds (AOR 1.31, 95% CI [1.10-1.56], p<0.01).

#### Differences by behaviour and demographics – adolescents

Logistic regression analyses indicated differences by age for open air playparks (AOR 1.10, 95% CI [1.05-1.15], p<0.001), school grounds (AOR 1.11, 95% CI [1.06-1.16], p<0.001) and public transport (AOR 1.08, 95% CI [1.03-1.13], p<0.01) with older participants more likely to doubt compliance (Supplementary Table 6b).

## DISCUSSION

Among adults and adolescents in this study, there was near consensus that vaping on public transport and on school grounds is unacceptable. More than half viewed vaping inside pubs, nightclubs and at hospital entrances as unacceptable. A minority of adults thought that vaping at open air playparks is unacceptable. A similar pattern was found for views on where vaping should not be allowed. The vast majority of adults and adolescents believed that vaping should not be allowed on public transport or on school grounds. Over half believed vaping should not be allowed inside pubs, nightclubs, and at hospital entrances. The most popular view among adults was that vaping in open air playparks should be allowed. Overall, adults and adolescents perceived likely compliance with a ban on vaping on public transport, on school grounds, and inside pubs but did not believe that people would obey a ban on vaping inside nightclubs, outside hospital entrances, and at open air playparks.

Responses varied with respondent characteristics. Adults who currently vaped and/or smoked, and adolescents who currently vaped, were more likely than those not currently vaping or smoking to consider it acceptable to vape, and more likely to believe that vaping should be allowed, in all six places proposed. It is well established in research assessing support for public health policies that acceptability varies according to participants’ behaviour. Those directly affected by a policy are generally more sceptical and less likely to support it.^22^ Just as people who do not, or used to, smoke are more likely than people who do smoke to support tobacco control interventions,^22^ people who do not vape are more likely than those that do to support vape-free policies.^28,31^ Here, we also found that people who smoke and/or vape were more likely, than those who do not, to agree that vaping should be allowed in public places. Our findings show a clear gender difference with adult women less likely than men to consider it acceptable to vape and that vaping should be allowed in all proposed places. This finding aligns with other studies demonstrating that women are more likely than men to support more intrusive policy measures for tobacco and alcohol.^22^

Sample differences were found for perceived compliance. For example, adults who vaped and/or smoked were less likely than non-users to doubt compliance with a ban on vaping at open air playparks and on school grounds. Dual users were less likely to doubt compliance outside hospital entrances. Older adolescents were more likely to doubt compliance at open air playparks, on school grounds and on public transport. One possible explanation is that views on compliance may reflect existing awareness of, and exposure to, the reality of these environments. For example, older adolescents may have observed people vaping in places where voluntary prohibitions are already in place. Future in-depth qualitative research may provide an understanding of what underpins these differing views.

The findings have implications for policymakers. Given our adult sample was limited to current and former nicotine users, it is plausible that among the general UK adult population, levels of unacceptability of, and support for, banning vaping in public places would likely be higher. Population-based surveys of other vaping and novel tobacco policies have demonstrated higher levels of public support among the general Great Britain population compared with key behavioural subgroups.^32,33^ Nonetheless, should policymakers decide to ban vaping in public places, our findings indicate which audiences may benefit from targeted communication strategies to improve policy acceptability and support.^24,31^ Helping those most affected by vape-free policy understand the rationale behind it may contribute to more successful and sustained implementation and enforcement.^24,34^ For example, high compliance with indoor smoking bans was associated with greater support among people who smoked. Once implemented, smoking bans were largely self-enforcing,^34^ with policy support and behavioural norms increasing post-implementation.^35,36^ Support for vape-free policy may also increase over time among people who vape. However, our findings indicate where there may be greater opposition to bans in specific places. That both adults and adolescents considered vaping on public transport and outside school grounds unacceptable may indicate existing social norms around vaping in these places. Doubt on compliance with bans inside nightclubs, outside hospital entrances, and at open air playparks suggests people may need convincing about the practical feasibly of bans in these places (although outdoor areas of health care settings are excluded from current UK proposals for vape-free on the premise of protecting medically vulnerable people while still helping adult smokers quit).^4^

The study has limitations. The surveys used non-probability samples which have issues for generalisability and prevalence estimates.^37^ The adult sample is not representative of current or former nicotine users. It does, however, reflect a broad mix of adults with different nicotine use behaviours, whose views policymakers are likely to consider. Sampling weights were derived for the adolescent sample to better reflect the UK adolescent population. Logistic regression analyses present associations that were adjusted for sociodemographic factors. While the findings offer useful insight into the views of those within the UK, their generalisability to other jurisdictions is limited. The recruitment approach, online survey administration, and reliance on self-reported data, may have resulted in social desirability bias. We cannot guarantee that participants had privacy to disclose their real views or behaviours. Recruitment of adolescents below the age of 16 was via their parent on the YouGov panel. Although the survey may have been completed at home with others present, it is designed to be completed on any device. Completion on a mobile phone may have helped protect participants’ privacy. We also sought to minimise bias through careful survey design. The public involvement and cognitive testing stage increased confidence that questions were comprehensible and reflected how members of the public considered the issue of vape-free places. This also allowed consistency of measures between the adults and adolescents. While asking participants about perceived compliance is hypothetical and may not reflect how people would behave in real life, the study offers a timely, pragmatic, snapshot of public opinion to assist UK policymakers.

## Conclusion

The findings indicate support for some vape-free places among adolescents and adults, particularly on public transport and school grounds. Responses were mixed for vape-free hospital entrances, pubs, and nightclubs, and support was generally low for a ban at open air playparks. Given that participants doubted likely compliance in nightclubs, outside hospital entrances and at open air playparks, the UK public may need convincing of the feasibility of banning vaping in these places.

## ACKNOWLEDGMENTS

The authors thank YouGov for delivery and management of the survey fieldwork, Jacqui Whyte for facilitating recruitment for the public involvement groups and cognitive interviews, Kathryn Angus for assisting with references, and all the participants who took part in the survey development phase or the survey,

## DATA AVAILABILITY

Participant consent was not obtained for data sharing beyond the scope of the original study.

