## Supplementary table 1a for "Adult current and former nicotine users’ and adolescents’ views on vape-free places: findings from UK cross-sectional surveys"

**Table S1a: Perceptions of acceptability of vaping in different locations: Adults**

| **Location** | **n** | **%** | **95% CI** |
| --- | --- | --- | --- |
| **Open air playparks** |  |  |  |
| 1 – Very acceptable | 601 | 21.1% | [19.6% - 22.6%] |
| 2 | 462 | 16.2% | [14.9% - 17.5%] |
| 3 | 596 | 20.9% | [19.4% - 22.4%] |
| 4 | 455 | 16.0% | [14.6% - 17.4%] |
| 5 – Very unacceptable | 703 | 24.7% | [23.1% - 26.3%] |
| Don’t Know | 34 | 1.2% | [0.8% - 1.6%] |
| Net: Acceptable | 1063 | 37.3% | [35.6% - 39.1%] |
| Net: Unacceptable | 1158 | 40.6% | [38.9% - 42.4%] |
| **Public transport** |  |  |  |
| 1 – Very acceptable | 106 | 3.7% | [3.0% - 4.4%] |
| 2 | 89 | 3.1% | [2.5% - 3.8%] |
| 3 | 190 | 6.7% | [5.8% - 7.5%] |
| 4 | 415 | 14.6% | [13.3% - 15.8%] |
| 5 – Very unacceptable | 2034 | 71.3% | [69.8% - 73.1%] |
| Don’t Know | 17 | 0.6% | [0.3% - 0.9%] |
| Net: Acceptable | 195 | 6.8% | [5.9% - 7.8%] |
| Net: Unacceptable | 2449 | 85.9% | [84.5% - 87.1%] |
| **Hospital entrances (outdoors)** |  |  |  |
| 1 – Very acceptable | 294 | 10.3% | [9.3% - 11.4%] |
| 2 | 302 | 10.6% | [9.4% - 11.8%] |
| 3 | 545 | 19.1% | [17.6% - 20.6%] |
| 4 | 524 | 18.4% | [16.9% - 19.8%] |
| 5 – Very unacceptable | 1163 | 40.8% | [38.9% - 42.6%] |
| Don’t Know | 23 | 0.8% | [0.5% - 1.2%] |
| Net: Acceptable | 596 | 20.9% | [19.4% - 22.4%] |
| Net: Unacceptable | 1687 | 59.2% | [57.3% - 61.0%] |
| *Base: All adults (N=2,851)* |  |  |  |

*Due to rounding, composites of ‘Unacceptable’ and ‘Acceptable’ may not appear to exactly add to the total of the individual items.*

**Table S1a cont’d: Perceptions of acceptability of vaping in different locations: Adults**

| **Location** | **n** | **%** | **95% CI** |
| --- | --- | --- | --- |
| **Inside nightclubs** |  |  |  |
| 1 – Very acceptable | 463 | 16.2% | [14.9% - 17.6%] |
| 2 | 389 | 13.6% | [12.4% - 14.9%] |
| 3 | 442 | 15.5% | [14.2% - 16.8%] |
| 4 | 396 | 13.9% | [12.6% - 15.2%] |
| 5 – Very unacceptable | 1102 | 38.7% | [36.9% - 40.5%] |
| Don’t Know | 59 | 2.1% | [1.5% - 2.6%] |
| Net: Acceptable | 852 | 29.9% | [28.2% - 31.6%] |
| Net: Unacceptable | 1498 | 52.5% | [50.8% - 54.4%] |
| **Pubs (indoors)** |  |  |  |
| 1 – Very acceptable | 375 | 13.2% | [11.9% - 14.4%] |
| 2 | 364 | 12.8% | [11.6% - 14.0%] |
| 3 | 432 | 15.2% | [13.9% - 16.5%] |
| 4 | 456 | 16.0% | [14.6% - 17.4%] |
| 5 – Very unacceptable | 1192 | 41.8% | [40.0% - 43.7%] |
| Don’t Know | 32 | 1.1% | [0.7% - 1.5%] |
| Net: Acceptable | 739 | 25.9% | [24.5% - 27.5%] |
| Net: Unacceptable | 1648 | 57.8% | [55.9% - 59.6%] |
| **On school grounds (outdoors)** |  |  |  |
| 1 – Very acceptable | 144 | 5.1% | [4.3% - 5.9%] |
| 2 | 92 | 3.2% | [2.6% - 3.9%] |
| 3 | 208 | 7.3% | [6.3% - 8.2%] |
| 4 | 367 | 12.9% | [11.6% - 14.1%] |
| 5 – Very unacceptable | 2017 | 70.7% | [69.1% - 72.5%] |
| Don’t Know | 23 | 0.8% | [0.5% - 1.1%] |
| Net: Acceptable | 236 | 8.3% | [7.3% - 9.3%] |
| Net: Unacceptable | 2384 | 83.6% | [82.3% - 85.0%] |

*Base: All adults (N=2,851)*

*Due to rounding, composites of ‘Unacceptable’ and ‘Acceptable’ may not appear to exactly add to the total of the individual items.*
