## Supplementary table 1b for "Adult current and former nicotine users’ and adolescents’ views on vape-free places: findings from UK cross-sectional surveys"

**Table S1b: Perceptions of acceptability of vaping in different locations: Adolescents**

| **Location** | **n** | **%** | **95% CI** |
| --- | --- | --- | --- |
| **Open air playparks** |  |  |  |
| 1 – Very acceptable | 130 | 6.1% | [5.1% - 7.2%] |
| 2 | 213 | 10.0% | [8.8% - 11.3%] |
| 3 | 360 | 17.0% | [15.4% - 18.6%] |
| 4 | 376 | 17.7% | [16.1% - 19.3%] |
| 5 – Very unacceptable | 970 | 45.7% | [43.6% - 47.8%] |
| Don’t Know | 73 | 3.5% | [2.7% - 4.2%] |
| Net: Acceptable | 343 | 16.2% | [14.6% -17.7%] |
| Net: Unacceptable | 1346 | 63.4% | [61.4% - 65.5%] |
| **Public transport** |  |  |  |
| 1 – Very acceptable | 38 | 1.8% | [1.2% - 2.3%] |
| 2 | 30 | 1.4% | [0.9% - 1.9%] |
| 3 | 88 | 4.1% | [3.3% - 5.0%] |
| 4 | 206 | 9.7% | [8.5% - 11.0%] |
| 5 – Very unacceptable | 1730 | 81.5% | [79.9% - 83.2%] |
| Don’t Know | 30 | 1.4% | [0.9% - 1.9%] |
| Net: Acceptable | 68 | 3.2% | [2.5% - 4.0%] |
| Net: Unacceptable | 1936 | 91.2% | [90.0% - 92.4%] |
| **Hospital entrances (outdoors)** |  |  |  |
| 1 – Very acceptable | 58 | 2.7% | [2.0% - 3.4%] |
| 2 | 53 | 2.5% | [1.8% - 3.2%] |
| 3 | 160 | 7.5% | [6.4% - 8.6%] |
| 4 | 251 | 11.8% | [10.5% - 13.2%] |
| 5 – Very unacceptable | 1547 | 72.9% | [71.0% - 74.8%] |
| Don’t Know | 54 | 2.5% | [1.9% - 3.2%] |
| Net: Acceptable | 111 | 5.2% | [4.3% - 6.2%] |
| Net: Unacceptable | 1798 | 84.7% | [83.2% - 86.3%] |
| *Base: All adolescents, weighted (N=2,123)* |  |  |  |

*Due to weighting and rounding, composites of ‘Unacceptable’ and ‘Acceptable’ may not appear to exactly add to the total of the individual items.*

**Table S1b cont’d: Perceptions of acceptability of vaping in different locations: Adolescents**

| **Location** | **n** | **%** | **95% CI** |
| --- | --- | --- | --- |
| **Inside nightclubs** |  |  |  |
| 1 – Very acceptable | 127 | 6.0% | [5.0% - 7.0%] |
| 2 | 170 | 8.0% | [6.8% - 9.2%] |
| 3 | 266 | 12.5% | [11.1% - 14.0%] |
| 4 | 253 | 11.9% | [10.6% - 13.3%] |
| 5 – Very unacceptable | 1087 | 51.2% | [49.1% - 53.4%] |
| Don’t Know | 219 | 10.3% | [9.0% - 11.6%] |
| Net: Acceptable | 297 | 14.0% | [12.5% - 15.5%] |
| Net: Unacceptable | 1341 | 63.2% | [61.1% - 65.2%] |
| **Pubs (indoors)** |  |  |  |
| 1 – Very acceptable | 102 | 4.8% | [3.9% - 5.7%] |
| 2 | 129 | 6.1% | [5.1% - 7.1%] |
| 3 | 231 | 10.9% | [9.5% - 12.2%] |
| 4 | 264 | 12.4% | [11.0% - 13.8%] |
| 5 – Very unacceptable | 1275 | 60.0% | [58.0% - 62.1%] |
| Don’t Know | 123 | 5.8% | [4.8% - 6.8%] |
| Net: Acceptable | 231 | 10.9% | [9.5% - 12.2%] |
| Net: Unacceptable | 1538 | 72.5% | [70.6% - 74.4%] |
| **On school grounds (outdoors)** |  |  |  |
| 1 – Very acceptable | 42 | 2.0% | [1.4% - 2.6%] |
| 2 | 38 | 1.8% | [1.2% - 2.3%] |
| 3 | 89 | 4.2% | [3.4% - 5.1%] |
| 4 | 170 | 8.0% | [6.8% - 9.2%] |
| 5 – Very unacceptable | 1750 | 82.5% | [80.8% - 84.1%] |
| Don’t Know | 34 | 1.6% | [1.1% - 2.1%] |
| Net: Acceptable | 80 | 3.7% | [2.9% - 4.6%] |
| Net: Unacceptable | 1920 | 90.5% | [89.2% - 91.7%] |

*Base: All adolescents, weighted (N=2,123)*

*Due to weighting and rounding, composites of ‘Unacceptable’ and ‘Acceptable’ may not appear to exactly add to the total of the individual items.*
