## Supplementary table 2a for "Adult current and former nicotine users’ and adolescents’ views on vape-free places: findings from UK cross-sectional surveys"

**Table S2a: Views on whether vaping should or should not be allowed in each location: Adults**

| **Location** | **n** | **%** | **95% CI** |
| --- | --- | --- | --- |
| **Open air playparks** |  |  |  |
| Should be allowed | 1409 | 49.4% | [47.6% - 51.3%] |
| Should not be allowed | 1177 | 41.3% | [39.5% - 43.1%] |
| Not sure | 265 | 9.3% | [8.2% - 10.4%] |
| **Public transport** |  |  |  |
| Should be allowed | 235 | 8.2% | [7.2% - 9.3%] |
| Should not be allowed | 2535 | 88.9% | [87.8% - 90.1%] |
| Not sure | 81 | 2.8% | [2.2% - 3.5%] |
| **Hospital entrances (outdoors)** |  |  |  |
| Should be allowed | 985 | 34.5% | [32.8% - 36.3%] |
| Should not be allowed | 1670 | 58.6% | [56.8% - 60.4%] |
| Not sure | 196 | 6.9% | [5.9% - 7.8%] |
| **Inside nightclubs** |  |  |  |
| Should be allowed | 1089 | 38.2% | [36.4% - 40.0%] |
| Should not be allowed | 1567 | 55.0% | [53.1% - 56.8%] |
| Not sure | 195 | 6.8% | [5.9% - 7.8%] |
| **Pubs (indoors)** |  |  |  |
| Should be allowed | 974 | 34.2% | [32.4% - 35.9%] |
| Should not be allowed | 1708 | 59.9% | [58.1% - 61.7%] |
| Not sure | 169 | 5.9% | [5.1% - 6.8%] |
| **On school grounds (outdoors)** |  |  |  |
| Should be allowed | 308 | 10.8% | [9.7% - 11.9%] |
| Should not be allowed | 2446 | 85.8% | [84.5% - 87.1%] |
| Not sure | 97 | 3.4% | [2.7% - 4.1%] |

*Base: All adults (N=2,851)*
