## Supplementary table 2b for "Adult current and former nicotine users’ and adolescents’ views on vape-free places: findings from UK cross-sectional surveys"

**Table S2b: Views on whether vaping should or should not be allowed in each location: Adolescents**

| **Location** | **n** | **%** | **95% CI** |
| --- | --- | --- | --- |
| **Open air playparks** |  |  |  |
| Should be allowed | 488 | 23.0% | [21.2% - 24.8%] |
| Should not be allowed | 1415 | 66.7% | [64.7% - 68.7%] |
| Not sure | 220 | 10.3% | [9.1% - 11.6%] |
| **Public transport** |  |  |  |
| Should be allowed | 73 | 3.4% | [2.7% - 4.2%] |
| Should not be allowed | 1992 | 93.8% | [92.8% - 94.9%] |
| Not sure | 58 | 2.7% | [2.0% - 3.4%] |
| **Hospital entrances (outdoors)** |  |  |  |
| Should be allowed | 178 | 8.4% | [7.2% - 9.6%] |
| Should not be allowed | 1834 | 86.4% | [85.0% - 87.9%] |
| Not sure | 110 | 5.2% | [4.2% - 6.1%] |
| **Inside nightclubs** |  |  |  |
| Should be allowed | 376 | 17.7% | [16.1% - 19.3%] |
| Should not be allowed | 1475 | 69.5% | [67.5% - 71.4%] |
| Not sure | 272 | 12.8% | [11.4% - 14.2%] |
| **Pubs (indoors)** |  |  |  |
| Should be allowed | 316 | 14.9% | [13.4% - 16.4%] |
| Should not be allowed | 1622 | 76.4% | [74.6% - 78.2%] |
| Not sure | 185 | 8.7% | [7.5% - 9.9%] |
| **On school grounds (outdoors)** |  |  |  |
| Should be allowed | 81 | 3.8% | [3.0% - 4.6%] |
| Should not be allowed | 1986 | 93.5% | [92.5% - 94.6%] |
| Not sure | 56 | 2.6% | [2.0% - 3.3%] |

*Base: All adolescents weighted (N=2,123)*
