## Supplementary table 3a for "Adult current and former nicotine users’ and adolescents’ views on vape-free places: findings from UK cross-sectional surveys"

**Table S3a: Logistic regressions: perceptions of acceptability of vaping in each location: Adults**

|  |  | **a** | | | |  | **b** | | | | **c** | | | |
| --- | --- | --- | --- | --- | --- | --- | --- | --- | --- | --- | --- | --- | --- | --- |
| **Dependent variable:** |  | **Public transport** | | | |  | **On school grounds (outdoors)** | | | | **Pubs (indoors)** | | | |
|  |  | **1= Acceptable (score 1-2) (n=195)** | | | |  | **1= Acceptable (score 1-2) (n=236)** | | | | **1= Acceptable (score 1-2) (n=739)** | | | |
|  |  | **0= Neutral or unacceptable^#^ (n=2656)** | | | |  | **0= Neutral or unacceptable^#^ (n=2615)** | | | | **0= Neutral or unacceptable^#^ (n=2112)** | | | |
|  |  | **N** | **AOR*** | **95% CI** | **P** |  | **N** | **AOR*** | **95% CI** | **P** | **N** | **AOR*** | **95% CI** | **P** |
| **Gender** |  |  |  |  |  |  |  |  |  |  |  |  |  |  |
| Male |  | 1437 | Ref |  |  |  | 1437 | Ref |  |  | 1437 | Ref |  |  |
| Female |  | 1414 | 0.57 | [0.42-0.77] | <0.001 |  | 1414 | 0.63 | [0.48-0.83] | <0.001 | 1414 | 0.54 | [0.45-0.65] | <0.001 |
| **Age group** |  |  |  |  | 0.425 |  |  |  |  | 0.227 |  |  |  | 0.008 |
| 18-24 |  | 92 | Ref |  |  |  | 92 | Ref |  |  | 92 | Ref |  |  |
| 25-34 *v 18-24* |  | 548 | 1.23 | [0.50-3.00] | 0.654 |  | 548 | 0.94 | [0.46-1.93] | 0.863 | 548 | 1.13 | [0.69-1.86] | 0.630 |
| 35-44 *v 18-34* |  | 498 | 1.04 | [0.59-1.84] | 0.879 |  | 498 | 1.02 | [0.64-1.62] | 0.943 | 498 | 0.90 | [0.66-1.24] | 0.532 |
| 45-54 *v 18-44* |  | 399 | 0.96 | [0.57-1.60] | 0.861 |  | 399 | 0.83 | [0.53-1.28] | 0.396 | 399 | 1.05 | [0.79-1.41] | 0.716 |
| 55-64 *v 18-54* |  | 626 | 1.36 | [0.91-2.01] | 0.133 |  | 626 | 0.74 | [0.50-1.08] | 0.117 | 626 | 0.86 | [0.68-1.1] | 0.242 |
| 65+ *v 18-64* |  | 688 | 0.79 | [0.52-1.20] | 0.270 |  | 688 | 0.70 | [0.48-1.01] | 0.056 | 688 | 0.66 | [0.52-0.84] | <0.001 |
| **Social Grade** |  |  |  |  |  |  |  |  |  |  |  |  |  |  |
| ABC1 |  | 1979 | Ref |  |  |  | 1979 | Ref |  |  | 1979 | Ref |  |  |
| C2DE |  | 872 | 0.94 | [0.68-1.31] | 0.709 |  | 872 | 1.14 | [0.84-1.53] | 0.401 | 872 | 1.15 | [0.95-1.39] | 0.163 |
| **Country** |  |  |  |  | 0.896 |  |  |  |  | 0.595 |  |  |  | 0.689 |
| England |  | 2308 | Ref |  |  |  | 2308 | Ref |  |  | 2308 | Ref |  |  |
| Wales |  | 198 | 1.18 | [0.68-2.07] | 0.554 |  | 198 | 0.72 | [0.39-1.33] | 0.297 | 198 | 0.97 | [0.68-1.38] | 0.860 |
| Scotland |  | 258 | 0.89 | [0.52-1.52] | 0.669 |  | 258 | 0.79 | [0.48-1.31] | 0.363 | 258 | 0.91 | [0.66-1.24] | 0.542 |
| N. Ireland |  | 87 | 1.05 | [0.47-2.34] | 0.899 |  | 87 | 0.84 | [0.38-1.86] | 0.673 | 87 | 1.27 | [0.79-2.04] | 0.323 |
| **Current vaping and/or smoking status** |  |  |  |  | <0.001 |  |  |  |  | 0.002 |  |  |  | <0.001 |
| Does not currently vape or smoke |  | 482 | Ref |  |  |  | 482 | Ref |  |  | 482 | Ref |  |  |
| Dual use |  | 540 | 3.40 | [1.87-6.2] | <0.001 |  | 540 | 2.65 | [1.56-4.50] | <0.001 | 540 | 6.53 | [4.5-9.46] | <0.001 |
| Exclusively vapes |  | 1025 | 2.35 | [1.32-4.17] | 0.004 |  | 1025 | 1.77 | [1.06-2.96] | 0.028 | 1025 | 4.71 | [3.31-6.7] | <0.001 |
| Exclusively smokes |  | 804 | 2.12 | [1.17-3.82] | 0.013 |  | 804 | 2.26 | [1.35-3.78] | 0.002 | 804 | 2.33 | [1.61-3.37] | <0.001 |
| Test of model coefficients |  | χ^2^ =40.855 | | df=13 | P<0.001 |  | χ^2^ =41.89 | | df=13 | P<0.001 | χ^2^ =254.63 | | df=13 | P<0.001 |
| Naglekerke R |  | 0.036 |  |  |  |  | 0.034 |  |  |  | 0.13 |  |  |  |

*Base: All adults (N=2,851);* * adjusted for all other variables in the model, AOR, adjusted odds ratio; ref, reference category; 95% CI, 95% confidence interval. ^#^ includes ‘not sure’ responses.

**Table S3a Cont’d: Logistic regressions: perceptions of acceptability of vaping in each location: Adults**

|  |  | **d** | | | |  | **e** | | | | **f** | | | |
| --- | --- | --- | --- | --- | --- | --- | --- | --- | --- | --- | --- | --- | --- | --- |
| **Dependent variable:** |  | **Inside nightclubs** | | | |  | **Hospital entrances (outdoors)** | | | | **Open air playparks** | | | |
|  |  | **1= Acceptable (score 1-2) (n=852)** | | | |  | **1= Acceptable (score 1-2) (n=596)** | | | | **1= Acceptable (score 1-2) (n=1063)** | | | |
|  |  | **0= Neutral or unacceptable^#^ (n=1999)** | | | |  | **0= Neutral or unacceptable^#^ (n=2255)** | | | | **0= Neutral or unacceptable^#^ (n=1788)** | | | |
|  |  | **N** | **AOR*** | **95% CI** | **P** |  | **N** | **AOR*** | **95% CI** | **P** | **N** | **AOR*** | **95% CI** | **P** |
| **Gender** |  |  |  |  |  |  |  |  |  |  |  |  |  |  |
| Male |  | 1437 | Ref |  |  |  | 1437 | Ref |  |  | 1437 | Ref |  |  |
| Female |  | 1414 | 0.59 | [0.49-0.70] | <0.001 |  | 1414 | 0.70 | [0.58-0.84] | <0.001 | 1414 | 0.70 | [0.60-0.82] | <0.001 |
| **Age group** |  |  |  |  | <0.001 |  |  |  |  | 0.724 |  |  |  | 0.442 |
| 18-24 |  | 92 | Ref |  |  |  | 92 | Ref |  |  | 92 | Ref |  |  |
| 25-34 *v 18-24* |  | 548 | 0.52 | [0.33-0.83] | 0.006 |  | 548 | 0.95 | [0.56-1.6] | 0.842 | 548 | 1.24 | [0.77-1.98] | 0.370 |
| 35-44 *v 18-34* |  | 498 | 0.55 | [0.41-0.74] | <0.001 |  | 498 | 0.97 | [0.69-1.36] | 0.848 | 498 | 1.26 | [0.93-1.69] | 0.133 |
| 45-54 *v 18-44* |  | 399 | 0.58 | [0.44-0.76] | <0.001 |  | 399 | 0.82 | [0.59-1.12] | 0.208 | 399 | 0.99 | [0.76-1.3] | 0.945 |
| 55-64 *v 18-54* |  | 626 | 0.52 | [0.41-0.66] | <0.001 |  | 626 | 1.09 | [0.85-1.41] | 0.496 | 626 | 1.14 | [0.91-1.42] | 0.247 |
| 65+ *v 18-64* |  | 688 | 0.40 | [0.31-0.51] | <0.001 |  | 688 | 0.93 | [0.73-1.18] | 0.546 | 688 | 0.93 | [0.76-1.15] | 0.503 |
| **Social Grade** |  |  |  |  |  |  |  |  |  |  |  |  |  |  |
| ABC1 |  | 1979 | Ref |  |  |  | 1979 | Ref |  |  | 1979 | Ref |  |  |
| C2DE |  | 872 | 1.02 | [0.85-1.24] | 0.802 |  | 872 | 1.24 | [1.01-1.51] | 0.039 | 872 | 1.08 | [0.91-1.29] | 0.370 |
| **Country** |  |  |  |  | 0.102 |  |  |  |  | 0.228 |  |  |  | 0.392 |
| England |  | 2308 | Ref |  |  |  | 2308 | Ref |  |  | 2308 | Ref |  |  |
| Wales |  | 198 | 1.01 | [0.72-1.42] | 0.947 |  | 198 | 0.80 | [0.55-1.18] | 0.262 | 198 | 0.89 | [0.65-1.21] | 0.446 |
| Scotland |  | 258 | 0.67 | [0.48-0.92] | 0.014 |  | 258 | 0.72 | [0.51-1.02] | 0.066 | 258 | 0.80 | [0.61-1.06] | 0.119 |
| N. Ireland |  | 87 | 1.05 | [0.65-1.68] | 0.856 |  | 87 | 0.96 | [0.57-1.61] | 0.883 | 87 | 1.07 | [0.69-1.66] | 0.770 |
| **Current vaping and/or smoking status** |  |  |  |  | <0.001 |  |  |  |  | <0.001 |  |  |  | <0.001 |
| Does not currently vape or smoke |  | 482 | Ref |  |  |  | 482 | Ref |  |  | 482 | Ref |  |  |
| Dual use |  | 540 | 5.25 | [3.75-7.34] | <0.001 |  | 540 | 4.25 | [2.87-6.31] | <0.001 | 540 | 3.98 | [2.95-5.36] | <0.001 |
| Exclusively vapes |  | 1025 | 3.71 | [2.71-5.09] | <0.001 |  | 1025 | 3.91 | [2.7-5.66] | <0.001 | 1025 | 3.52 | [2.68-4.62] | <0.001 |
| Exclusively smokes |  | 804 | 1.90 | [1.36-2.65] | <0.001 |  | 804 | 3.08 | [2.11-4.51] | <0.001 | 804 | 2.73 | [2.06-3.61] | <0.001 |
| Test of model coefficients |  | χ^2^ =340.311 | | df=13 | P<0.001 |  | χ^2^ =105.157 | | df=13 | P<0.001 | χ^2^ =148.718 | | df=13 | P<0.001 |
| Naglekerke R |  | 0.16 |  |  |  |  | 0.056 |  |  |  | 0.069 |  |  |  |

*Base: All adults (N=2,851);* * adjusted for all other variables in the model, AOR, adjusted odds ratio; ref, reference category; 95% CI, 95% confidence interval. ^#^ includes ‘not sure’ responses.
