## Supplementary table 3b for "Adult current and former nicotine users’ and adolescents’ views on vape-free places: findings from UK cross-sectional surveys"

**Table S3b: Logistic regressions: perceptions of acceptability of vaping in each location: Adolescents**

|  |  | **a** | | | |  | **b** | | | | **c** | | | |
| --- | --- | --- | --- | --- | --- | --- | --- | --- | --- | --- | --- | --- | --- | --- |
| **Dependent variable:** |  | **Public transport** | | | |  | **On school grounds (outdoors)** | | | | **Pubs (indoors)** | | | |
|  |  | **1= Acceptable (score 1-2) (n=69)** | | | |  | **1= Acceptable (score 1-2) (n=78)** | | | | **1= Acceptable (score 1-2) (n=222)** | | | |
|  |  | **0= Neutral or unacceptable^#^ (n=2040)** | | | |  | **0= Neutral or unacceptable^#^ (n=2031)** | | | | **0= Neutral or unacceptable^#^ (n=1887)** | | | |
|  |  | **N** | **AOR*** | **95% CI** | **P** |  | **N** | **AOR*** | **95% CI** | **P** | **N** | **AOR*** | **95% CI** | **P** |
| **Gender** |  |  |  |  |  |  |  |  |  |  |  |  |  |  |
| Male |  | 1136 | Ref |  |  |  | 1136 | Ref |  |  | 1136 | Ref |  |  |
| Female |  | 973 | 1.15 | [0.70-1.89] | 0.575 |  | 973 | 1.28 | [0.80-2.05] | 0.294 | 973 | 0.62 | [0.46-0.83] | 0.001 |
| **Age group** |  |  | 0.99 | [0.87-1.13] | 0.913 |  |  | 1.00 | [0.89-1.13] | 0.988 |  | 0.95 | [0.88-1.02] | 0.168 |
| **Social Grade** |  |  |  |  |  |  |  |  |  |  |  |  |  |  |
| ABC1 |  | 1545 | Ref |  |  |  | 1545 | Ref |  |  | 1545 | Ref |  |  |
| C2DE |  | 564 | 0.74 | [0.41-1.33] | 0.319 |  | 564 | 1.28 | [0.78-2.11] | 0.324 | 564 | 0.89 | [0.64-1.24] | 0.495 |
| **Country** |  |  |  |  | 0.480 |  |  |  |  | 0.706 |  |  |  | 0.755 |
| England |  | 1742 | Ref |  |  |  | 1742 | Ref |  |  | 1742 | Ref |  |  |
| Wales |  | 193 | 1.54 | [0.73-3.25] | 0.257 |  | 193 | 1.13 | [0.52-2.46] | 0.755 | 193 | 1.24 | [0.78-1.99] | 0.364 |
| Scotland |  | 103 | 1.65 | [0.56-4.8] | 0.362 |  | 103 | 1.03 | [0.31-3.43] | 0.966 | 103 | 0.81 | [0.38-1.72] | 0.589 |
| N. Ireland |  | 71 | 0.52 | [0.07-3.87] | 0.519 |  | 71 | 1.89 | [0.65-5.52] | 0.246 | 71 | 1.00 | [0.44-2.27] | 0.992 |
| **Current vaping and/or smoking status** |  |  |  |  |  |  |  |  |  |  |  |  |  |  |
| Does not currently vape |  | 1966 | Ref |  |  |  | 1966 | Ref |  |  | 1966 | Ref |  |  |
| Currently vapes |  | 143 | 9.75 | [5.63-16.89] | <0.001 |  | 143 | 9.53 | [5.66-16.03] | <0.001 | 143 | 6.55 | [4.44-9.65] | <0.001 |
| Test of model coefficients |  | χ^2^ =57.590 | | df=7 | P<0.001 |  | χ^2^ =63.47 | | df=7 | P<0.001 | χ^2^ =92.42 | | df=7 | P<0.001 |
| Naglekerke R |  | 0.108 |  |  |  |  | 0.109 |  |  |  | 0.088 |  |  |  |

*Base: All adolescents, unweighted (N=2,109), missing cases (n=14) due to missing data on social grade (n=13) and current vaping (n=1)*

* adjusted for all other variables in the model, AOR, adjusted odds ratio; ref, reference category; 95% CI, 95% confidence interval. ^#^ includes ‘not sure’ responses.

**Table S3b Cont’d: Logistic regressions: perceptions of acceptability of vaping in each location: Adolescents**

|  |  | **d** | | | |  | **e** | | | | **f** | | | |
| --- | --- | --- | --- | --- | --- | --- | --- | --- | --- | --- | --- | --- | --- | --- |
| **Dependent variable:** |  | **Inside nightclubs** | | | |  | **Hospital entrances (outdoors)** | | | | **Open air playparks** | | | |
|  |  | **1= Acceptable (score 1-2) (n=283)** | | | |  | **1= Acceptable (score 1-2) (n=110)** | | | | **1= Acceptable (score 1-2) (n=337)** | | | |
|  |  | **0= Neutral or unacceptable^#^ (n=1826)** | | | |  | **0= Neutral or unacceptable^#^ (n=1999)** | | | | **0= Neutral or unacceptable^#^ (n=1772)** | | | |
|  |  | **N** | **AOR*** | **95% CI** | **P** |  | **N** | **AOR*** | **95% CI** | **P** | **N** | **AOR*** | **95% CI** | **P** |
| **Gender** |  |  |  |  |  |  |  |  |  |  |  |  |  |  |
| Male |  | 1136 | Ref |  |  |  | 1136 | Ref |  |  | 1136 | Ref |  |  |
| Female |  | 973 | 0.66 | [0.50-0.86] | 0.002 |  | 973 | 0.94 | [0.63-1.40] | 0.759 | 973 | 0.75 | [0.58-0.95] | 0.020 |
| **Age** |  |  | 0.95 | [0.89-1.02] | 0.178 |  |  | 1.04 | [0.94-1.15] | 0.481 |  | 1.08 | [1.02-1.15] | 0.011 |
| **Social Grade** |  |  |  |  |  |  |  |  |  |  |  |  |  |  |
| ABC1 |  | 1545 | Ref |  |  |  | 1545 | Ref |  |  | 1545 | Ref |  |  |
| C2DE |  | 564 | 0.73 | [0.54-1.00] | 0.048 |  | 564 | 0.91 | [0.58-1.42] | 0.672 | 564 | 1.06 | [0.81-1.39] | 0.678 |
| **Country** |  |  |  |  | 0.590 |  |  |  |  | 0.737 |  |  |  | 0.583 |
| England |  | 1742 | Ref |  |  |  | 1742 | Ref |  |  | 1742 | Ref |  |  |
| Wales |  | 193 | 1.02 | [0.65-1.60] | 0.940 |  | 193 | 1.37 | [0.74-2.55] | 0.314 | 193 | 0.73 | [0.46-1.15] | 0.176 |
| Scotland |  | 103 | 0.79 | [0.40-1.56] | 0.496 |  | 103 | 1.24 | [0.48-3.19] | 0.656 | 103 | 0.95 | [0.51-1.74] | 0.856 |
| N. Ireland |  | 71 | 1.48 | [0.77-2.87] | 0.241 |  | 71 | 1.27 | [0.44-3.62] | 0.660 | 71 | 1.08 | [0.57-2.08] | 0.808 |
| **Current vaping and/or smoking status** |  |  |  |  |  |  |  |  |  |  |  |  |  |  |
| Does not currently vape |  | 1966 | Ref |  |  |  | 1966 | Ref |  |  | 1966 | Ref |  |  |
| Currently vapes |  | 143 | 7.87 | [5.43-11.41] | <0.001 |  | 143 | 6.45 | [4.03-10.33] | <0.001 | 143 | 6.18 | [4.31-8.85] | <0.001 |
| Test of model coefficients |  | χ^2^ =125.34 | | df=7 | P<0.001 |  | χ^2^ =53.63 | | df=7 | P<0.001 | χ^2^ =117.984 | | df=7 | P<0.001 |
| Naglekerke R |  | 0.106 |  |  |  |  | 0.075 |  |  |  | 0.093 |  |  |  |

*Base: All adolescents, unweighted (N=2,109), missing cases (n=14) due to missing data on social grade (n=13) and current vaping (n=1)*

* adjusted for all other variables in the model, AOR, adjusted odds ratio; ref, reference category; 95% CI, 95% confidence interval. ^#^ includes ‘not sure’ responses.
