## Supplementary table 4a for "Adult current and former nicotine users’ and adolescents’ views on vape-free places: findings from UK cross-sectional surveys"

**Table S4a: Logistic regressions: views on whether vaping should be allowed in each location: Adults**

|  |  | **a** | | | |  | **b** | | | | **c** | | | |
| --- | --- | --- | --- | --- | --- | --- | --- | --- | --- | --- | --- | --- | --- | --- |
| **Dependent variable:** |  | **Public transport** | | | |  | **On school grounds (outdoors)** | | | | **Pubs (indoors)** | | | |
|  |  | **1= Should be allowed (n=235)** | | | |  | **1= Should be allowed (n=308)** | | | | **1= Should be allowed (n=974)** | | | |
|  |  | **0= Should not or not sure^#^ (n=2616)** | | | |  | **0= Should not or not sure^#^ (n=2543)** | | | | **0= Should not or not sure^#^ (n=1877)** | | | |
|  |  | **N** | **AOR*** | **95% CI** | **P** |  | **N** | **AOR*** | **95% CI** | **P** | **N** | **AOR*** | **95% CI** | **P** |
| **Gender** |  |  |  |  |  |  |  |  |  |  |  |  |  |  |
| Male |  | 1437 | Ref |  |  |  | 1437 | Ref |  |  | 1437 | Ref |  |  |
| Female |  | 1414 | 0.71 | [0.54-0.93] | 0.014 |  | 1414 | 0.62 | [0.49-0.8] | <0.001 | 1414 | 0.58 | [0.49-0.69] | <0.001 |
| **Age group** |  |  |  |  | 0.279 |  |  |  |  | 0.765 |  |  |  | 0.035 |
| 18-24 |  | 92 | Ref |  |  |  | 92 | Ref |  |  | 92 | Ref |  |  |
| 25-34 *v 18-24* |  | 548 | 0.91 | [0.41-2.02] | 0.818 |  | 548 | 0.99 | [0.5-1.97] | 0.985 | 548 | 0.82 | [0.51-1.3] | 0.395 |
| 35-44 *v 18-34* |  | 498 | 0.98 | [0.58-1.64] | 0.935 |  | 498 | 1.05 | [0.68-1.62] | 0.840 | 498 | 0.89 | [0.66-1.2] | 0.432 |
| 45-54 *v 18-44* |  | 399 | 1.18 | [0.75-1.86] | 0.467 |  | 399 | 0.75 | [0.5-1.15] | 0.189 | 399 | 0.97 | [0.74-1.27] | 0.807 |
| 55-64 *v 18-54* |  | 626 | 1.41 | [0.99-2.02] | 0.058 |  | 626 | 0.97 | [0.69-1.35] | 0.844 | 626 | 0.81 | [0.65-1.02] | 0.077 |
| 65+ *v 18-64* |  | 688 | 0.89 | [0.61-1.28] | 0.528 |  | 688 | 0.92 | [0.67-1.26] | 0.598 | 688 | 0.70 | [0.56-0.87] | 0.001 |
| **Social Grade** |  |  |  |  |  |  |  |  |  |  |  |  |  |  |
| ABC1 |  | 1979 | Ref |  |  |  | 1979 | Ref |  |  | 1979 | Ref |  |  |
| C2DE |  | 872 | 1.13 | [0.84-1.52] | 0.412 |  | 872 | 1.03 | [0.79-1.34] | 0.830 | 872 | 1.13 | [0.95-1.36] | 0.173 |
| **Country** |  |  |  |  | 0.577 |  |  |  |  | 0.459 |  |  |  | 0.261 |
| England |  | 2308 | Ref |  |  |  | 2308 | Ref |  |  | 2308 | Ref |  |  |
| Wales |  | 198 | 1.19 | [0.72-1.97] | 0.491 |  | 198 | 0.82 | [0.49-1.37] | 0.444 | 198 | 0.90 | [0.65-1.25] | 0.543 |
| Scotland |  | 258 | 0.75 | [0.44-1.26] | 0.274 |  | 258 | 1.30 | [0.88-1.91] | 0.182 | 258 | 0.79 | [0.59-1.06] | 0.120 |
| N. Ireland |  | 87 | 1.14 | [0.56-2.33] | 0.713 |  | 87 | 0.99 | [0.50-1.94] | 0.971 | 87 | 1.27 | [0.81-2.00] | 0.299 |
| **Current vaping and/or smoking status** |  |  |  |  | <0.001 |  |  |  |  | <0.001 |  |  |  | <0.001 |
| Does not currently vape or smoke |  | 482 | Ref |  |  |  | 482 | Ref |  |  | 482 | Ref |  |  |
| Dual use |  | 540 | 3.74 | [2.13-6.56] | <0.001 |  | 540 | 2.63 | [1.63-4.23] | <0.001 | 540 | 5.98 | [4.32-8.28] | <0.001 |
| Exclusively vapes |  | 1025 | 2.81 | [1.64-4.81] | <0.001 |  | 1025 | 2.15 | [1.37-3.37] | <0.001 | 1025 | 5.08 | [3.76-6.87] | <0.001 |
| Exclusively smokes |  | 804 | 2.12 | [1.21-3.7] | 0.008 |  | 804 | 2.39 | [1.52-3.75] | <0.001 | 804 | 2.23 | [1.63-3.06] | <0.001 |
| Test of model coefficients |  | χ^2^ =43.560 | | df=13 | P<0.001 |  | χ^2^ =42.593 | | df=13 | P<0.001 | χ^2^ =292.782 | | df=13 | P<0.001 |
| Naglekerke R |  | 0.035 |  |  |  |  | 0.030 |  |  |  | 0.135 |  |  |  |

*Base: All adults (N=2,851);* * adjusted for all other variables in the model, AOR, adjusted odds ratio; ref, reference category; 95% CI, 95% confidence interval.

**Table S4a Cont’d: Logistic regressions: views on whether vaping should be allowed in each location: Adults**

|  |  | **d** | | | |  | **e** | | | | **f** | | | |
| --- | --- | --- | --- | --- | --- | --- | --- | --- | --- | --- | --- | --- | --- | --- |
| **Dependent variable:** |  | **Inside nightclubs** | | | |  | **Hospital entrances (outdoors)** | | | | **Open air playparks** | | | |
|  |  | **1= Should be allowed (n=1089)** | | | |  | **1= Should be allowed (n=985)** | | | | **1= Should be allowed (n=1409)** | | | |
|  |  | **0= Should not or not sure^#^ (n=1762)** | | | |  | **0= Should not or not sure^#^ (n=1866)** | | | | **0= Should not or not sure^#^ (n=1442)** | | | |
|  |  | **N** | **AOR*** | **95% CI** | **P** |  | **N** | **AOR*** | **95% CI** | **P** | **N** | **AOR*** | **95% CI** | **P** |
| **Gender** |  |  |  |  |  |  |  |  |  |  |  |  |  |  |
| Male |  | 1437 | Ref |  |  |  | 1437 | Ref |  |  | 1437 | Ref |  |  |
| Female |  | 1414 | 0.60 | [0.51-0.7] | <0.001 |  | 1414 | 0.82 | [0.7-0.97] | 0.018 | 1414 | 0.69 | [0.59-0.8] | <0.001 |
| **Age group** |  |  |  |  | <0.001 |  |  |  |  | 0.408 |  |  |  | 0.527 |
| 18-24 |  | 92 | Ref |  |  |  | 92 | Ref |  |  | 92 | Ref |  |  |
| 25-34 *v 18-24* |  | 548 | 0.34 | [0.2-0.56] | <0.001 |  | 548 | 0.86 | [0.54-1.37] | 0.514 | 548 | 0.77 | [0.49-1.2] | 0.248 |
| 35-44 *v 18-34* |  | 498 | 0.47 | [0.34-0.65] | <0.001 |  | 498 | 1.00 | [0.74-1.34] | 0.974 | 498 | 1.01 | [0.76-1.35] | 0.953 |
| 45-54 *v 18-44* |  | 399 | 0.66 | [0.5-0.87] | 0.004 |  | 399 | 1.11 | [0.85-1.46] | 0.428 | 399 | 0.92 | [0.71-1.2] | 0.549 |
| 55-64 *v 18-54* |  | 626 | 0.49 | [0.39-0.62] | <0.001 |  | 626 | 1.13 | [0.9-1.4] | 0.290 | 626 | 1.11 | [0.9-1.37] | 0.346 |
| 65+ *v 18-64* |  | 688 | 0.42 | [0.33-0.52] | <0.001 |  | 688 | 1.13 | [0.92-1.39] | 0.250 | 688 | 0.99 | [0.82-1.21] | 0.940 |
| **Social Grade** |  |  |  |  |  |  |  |  |  |  |  |  |  |  |
| ABC1 |  | 1979 | Ref |  |  |  | 1979 | Ref |  |  | 1979 | Ref |  |  |
| C2DE |  | 872 | 1.05 | [0.88-1.26] | 0.590 |  | 872 | 1.21 | [1.02-1.44] | 0.029 | 872 | 1.05 | [0.89-1.24] | 0.555 |
| **Country** |  |  |  |  | 0.048 |  |  |  |  | 0.493 |  |  |  | 0.793 |
| England |  | 2308 | Ref |  |  |  | 2308 | Ref |  |  | 2308 | Ref |  |  |
| Wales |  | 198 | 1.03 | [0.75-1.41] | 0.872 |  | 198 | 1.04 | [0.77-1.42] | 0.783 | 198 | 0.92 | [0.68-1.24] | 0.588 |
| Scotland |  | 258 | 0.66 | [0.49-0.9] | 0.007 |  | 258 | 0.87 | [0.66-1.16] | 0.340 | 258 | 1.08 | [0.83-1.41] | 0.578 |
| N. Ireland |  | 87 | 1.14 | [0.72-1.8] | 0.569 |  | 87 | 0.75 | [0.47-1.2] | 0.229 | 87 | 1.15 | [0.74-1.78] | 0.541 |
| **Current vaping and/or smoking status** |  |  |  |  | <0.001 |  |  |  |  | <0.001 |  |  |  | <0.001 |
| Does not currently vape or smoke |  | 482 | Ref |  |  |  | 482 | Ref |  |  | 482 | Ref |  |  |
| Dual use |  | 540 | 4.66 | [3.45-6.3] | <0.001 |  | 540 | 4.27 | [3.12-5.84] | <0.001 | 540 | 3.46 | [2.64-4.52] | <0.001 |
| Exclusively vapes |  | 1025 | 3.90 | [2.96-5.13] | <0.001 |  | 1025 | 4.13 | [3.09-5.5] | <0.001 | 1025 | 2.97 | [2.34-3.77] | <0.001 |
| Exclusively smokes |  | 804 | 1.70 | [1.27-2.27] | <0.001 |  | 804 | 3.14 | [2.34-4.21] | <0.001 | 804 | 2.32 | [1.82-2.96] | <0.001 |
| Test of model coefficients |  | χ^2^ =372.781 | | df=13 | P<0.001 |  | χ^2^ =139.642 | | df=13 | P<0.001 | χ^2^ =137.915 | | df=13 | P<0.001 |
| Naglekerke R |  | 0.17 |  |  |  |  | 0.066 |  |  |  | 0.063 |  |  |  |

*Base: All adults (N=2,851);* * adjusted for all other variables in the model, AOR, adjusted odds ratio; ref, reference category; 95% CI, 95% confidence interval.
