## Supplementary table 4b for "Adult current and former nicotine users’ and adolescents’ views on vape-free places: findings from UK cross-sectional surveys"

**Table S4b: Logistic regressions: views on whether vaping should be allowed in each location: Adolescents**

|  |  | **a** | | | |  | **b** | | | | **c** | | | |
| --- | --- | --- | --- | --- | --- | --- | --- | --- | --- | --- | --- | --- | --- | --- |
| **Dependent variable:** |  | **Public transport** | | | |  | **On school grounds (outdoors)** | | | | **Pubs (indoors)** | | | |
|  |  | **1= Should be allowed (n=74)** | | | |  | **1= Should be allowed (n=78)** | | | | **1= Should be allowed (n=308)** | | | |
|  |  | **0= Should not or not sure (n=2035)** | | | |  | **0= Should not or not sure (n=2031)** | | | | **0= Should not or not sure (n=1801)** | | | |
|  |  | **N** | **AOR*** | **95% CI** | **P** |  | **N** | **AOR*** | **95% CI** | **P** | **N** | **AOR*** | **95% CI** | **P** |
| **Gender** |  |  |  |  |  |  |  |  |  |  |  |  |  |  |
| Male |  | 1136 | Ref |  |  |  | 1136 | Ref |  |  | 1136 | Ref |  |  |
| Female |  | 973 | 0.54 | [0.32-0.90] | 0.019 |  | 973 | 0.78 | [0.48-1.26] | 0.313 | 973 | 0.57 | [0.44-0.74] | <0.001 |
| **Age group** |  |  | 0.94 | [0.83-1.07] | 0.382 |  |  | 0.98 | [0.87-1.11] | 0.785 |  | 0.99 | [0.93-1.06] | 0.871 |
| **Social Grade** |  |  |  |  |  |  |  |  |  |  |  |  |  |  |
| ABC1 |  | 1545 | Ref |  |  |  | 1545 |  |  |  | 1545 | Ref |  |  |
| C2DE |  | 564 | 1.00 | [0.58-1.72] | 0.998 |  | 564 | 0.94 | [0.55-1.59] | 0.807 | 564 | 0.92 | [0.69-1.22] | 0.567 |
| **Country** |  |  |  |  | 0.019 |  |  |  |  | 0.634 |  |  |  | 0.441 |
| England |  | 1742 | Ref |  |  |  | 1742 |  |  |  | 1742 | Ref |  |  |
| Wales |  | 193 | 2.56 | [1.34-4.90] | 0.004 |  | 193 | 0.78 | [0.33-1.87] | 0.583 | 193 | 1.38 | [0.92-2.06] | 0.123 |
| Scotland |  | 103 | 0.41 | [0.05-3.05] | 0.383 |  | 103 | 0.32 | [0.04-2.35] | 0.261 | 103 | 1.23 | [0.69-2.19] | 0.488 |
| N. Ireland |  | 71 | 0.46 | [0.06-3.51] | 0.450 |  | 71 | 1.26 | [0.37-4.26] | 0.708 | 71 | 1.05 | [0.52-2.12] | 0.893 |
| **Current vaping and/or smoking status** |  |  |  |  |  |  |  |  |  |  |  |  |  |  |
| Does not currently vape |  | 1966 | Ref |  |  |  | 1966 |  |  |  | 1966 | Ref |  |  |
| Currently vapes |  | 143 | 14.66 | [8.66-24.83] | <0.001 |  | 143 | 9.83 | [5.87-16.46] | <0.001 | 143 | 6.39 | [4.43-9.22] | <0.001 |
| Test of model coefficients |  | χ^2^ =103.755 | | df=7 | P<0.001 |  | χ^2^ =68.220 | | df=7 | P<0.001 | χ^2^ =114.604 | | df=7 | P<0.001 |
| Naglekerke R |  | 0.183 |  |  |  |  | 0.117 |  |  |  | 0.094 |  |  |  |

*Base: All adolescents, unweighted (N=2,109), missing cases (n=14) due to missing data on social grade (n=13) and current vaping (n=1)*

* adjusted for all other variables in the model, AOR, adjusted odds ratio; ref, reference category; 95% CI, 95% confidence interval.

**Table S4b Cont’d: Logistic regressions: views on whether vaping should be allowed in each location: Adolescents**

|  |  | **d** | | | |  | **e** | | | | **f** | | | |
| --- | --- | --- | --- | --- | --- | --- | --- | --- | --- | --- | --- | --- | --- | --- |
| **Dependent variable:** |  | **Inside nightclubs** | | | |  | **Hospital entrances (outdoors)** | | | | **Open air playparks** | | | |
|  |  | **1= Should be allowed (n=360)** | | | |  | **1= Should be allowed (n=176)** | | | | **1= Should be allowed (n=483)** | | | |
|  |  | **0= Should not or not sure (n=1749)** | | | |  | **0= Should not or not sure (n=1933)** | | | | **0= Should not or not sure (n=1626)** | | | |
|  |  | **N** | **AOR*** | **95% CI** | **P** |  | **N** | **AOR*** | **95% CI** | **P** | **N** | **AOR*** | **95% CI** | **P** |
| **Gender** |  |  |  |  |  |  |  |  |  |  |  |  |  |  |
| Male |  | 1136 | Ref |  |  |  | 1136 | Ref |  |  | 1136 | Ref |  |  |
| Female |  | 973 | 0.61 | [0.48-0.78] | <0.001 |  | 973 | 0.82 | [0.59-1.13] | 0.216 | 973 | 0.71 | [0.57-0.87] | 0.001 |
| **Age** |  |  | 0.98 | [0.92-1.04] | 0.443 |  |  | 1.05 | [0.97-1.14] | 0.228 |  | 1.06 | [1.00-1.12] | 0.044 |
| **Social Grade** |  |  |  |  |  |  |  |  |  |  |  |  |  |  |
| ABC1 |  | 1545 | Ref |  |  |  | 1545 | Ref |  |  | 1545 | Ref |  |  |
| C2DE |  | 564 | 0.84 | [0.64-1.11] | 0.216 |  | 564 | 1.05 | [0.74-1.50] | 0.768 | 564 | 1.20 | [0.95-1.52] | 0.121 |
| **Country** |  |  |  |  | 0.631 |  |  |  |  | 0.539 |  |  |  | 0.450 |
| England |  | 1742 | Ref |  |  |  | 1742 | Ref |  |  | 1742 | Ref |  |  |
| Wales |  | 193 | 0.90 | [0.59-1.37] | 0.627 |  | 193 | 0.94 | [0.54-1.62] | 0.816 | 193 | 0.80 | [0.55-1.17] | 0.246 |
| Scotland |  | 103 | 0.91 | [0.51-1.61] | 0.747 |  | 103 | 0.52 | [0.19-1.45] | 0.212 | 103 | 0.72 | [0.41-1.25] | 0.237 |
| N. Ireland |  | 71 | 0.62 | [0.29-1.34] | 0.223 |  | 71 | 0.66 | [0.23-1.84] | 0.424 | 71 | 1.03 | [0.58-1.83] | 0.910 |
| **Current vaping and/or smoking status** |  |  |  |  |  |  |  |  |  |  |  |  |  |  |
| Does not currently vape |  | 1966 | Ref |  |  |  | 1966 | Ref |  |  | 1966 | Ref |  |  |
| Currently vapes |  | 143 | 6.24 | [4.34-8.95] | <0.001 |  | 143 | 4.80 | [3.17-7.26] | <0.001 | 143 | 5.35 | [3.74-7.64] | <0.001 |
| Test of model coefficients |  | χ^2^ =115.223 | | df=7 | P<0.001 |  | χ^2^ =58.592 | | df=7 | P<0.001 | χ^2^ =115.763 | | df=7 | P<0.001 |
| Naglekerke R |  | 0.089 |  |  |  |  | 0.063 |  |  |  | 0.081 |  |  |  |

*Base: All adolescents, unweighted (N=2,109), missing cases (n=14) due to missing data on social grade (n=13) and current vaping (n=1)*

* adjusted for all other variables in the model, AOR, adjusted odds ratio; ref, reference category; 95% CI, 95% confidence interval.
