## Supplementary table 5a for "Adult current and former nicotine users’ and adolescents’ views on vape-free places: findings from UK cross-sectional surveys"

**Table S5a: Perceived likelihood of people obeying rules in each location if vaping was banned: Adults**

| **Location** | **n** | **%** | **95% CI** |
| --- | --- | --- | --- |
| **Open air playparks** |  |  |  |
| Very likely to obey | 277 | 9.7% | [8.6% - 10.8%] |
| Quite likely to obey | 456 | 16.0% | [14.6% - 17.3%] |
| Quite unlikely to obey | 920 | 32.3% | [30.6% - 34.0%] |
| Very unlikely to obey | 1080 | 37.9% | [36.1% - 39.7%] |
| Not sure | 118 | 4.1% | [3.4% - 4.9%] |
| Net: Likely | 733 | 25.7% | [24.1% - 27.3%] |
| Net: Unlikely | 2000 | 70.2% | [68.5% - 71.5%] |
| **Public transport** |  |  |  |
| Very likely to obey | 990 | 34.7% | [33.0% - 36.5%] |
| Quite likely to obey | 1079 | 37.8% | [36.1% - 39.6%] |
| Quite unlikely to obey | 438 | 15.4% | [14.0% - 16.7%] |
| Very unlikely to obey | 243 | 8.5% | [7.5% - 9.5%] |
| Don’t Know | 101 | 3.5% | [2.9% - 4.2%] |
| Net: Likely | 2069 | 72.6% | [70.9% - 74.2%] |
| Net: Unlikely | 681 | 23.9% | [22.3% - 25.5%] |
| **Hospital entrances (outdoors)** |  |  |  |
| Very likely to obey | 418 | 14.7% | [13.4% - 16.0%] |
| Quite likely to obey | 593 | 20.8% | [19.3% - 22.3%] |
| Quite unlikely to obey | 937 | 32.9% | [31.2% - 34.6%] |
| Very unlikely to obey | 787 | 27.6% | [26.0% - 29.2%] |
| Don’t Know | 116 | 4.1% | [3.3% - 4.8%] |
| Net: Likely | 1011 | 35.5% | [33.7% - 37.2%] |
| Net: Unlikely | 1724 | 60.5% | [58.7% - 62.3%] |
| *Base: All adults (N=2,851)* |  |  |  |

**Table S5a cont’d: Perceived likelihood of people obeying rules in each location if vaping was banned: Adults**

| **Location** | **n** | **%** | **95% CI** |
| --- | --- | --- | --- |
| **Inside nightclubs** |  |  |  |
| Very likely to obey | 514 | 18.0% | [16.6% - 19.4%] |
| Quite likely to obey | 690 | 24.2% | [22.6% - 25.8%] |
| Quite unlikely to obey | 771 | 27.0% | [25.4% - 28.7%] |
| Very unlikely to obey | 743 | 26.1% | [24.4% - 27.7%] |
| Don’t Know | 133 | 4.7% | [3.9% - 5.4%] |
| Net: Likely | 1204 | 42.2% | [40.4% - 44.0%] |
| Net: Unlikely | 1514 | 53.1% | [51.3% - 54.9%] |
| **Pubs (indoors)** |  |  |  |
| Very likely to obey | 694 | 24.3% | [22.8% - 25.9%] |
| Quite likely to obey | 952 | 33.4% | [31.7% - 35.1%] |
| Quite unlikely to obey | 654 | 22.9% | [21.4% - 24.5%] |
| Very unlikely to obey | 447 | 15.7% | [14.3% - 17.0%] |
| Don’t Know | 104 | 3.6% | [3.0% - 4.3%] |
| Net: Likely | 1646 | 57.7% | [55.9% - 59.5%] |
| Net: Unlikely | 1101 | 38.6% | [36.8% - 40.4%] |
| **On school grounds (outdoors)** |  |  |  |
| Very likely to obey | 839 | 29.4% | [27.8% - 31.1%] |
| Quite likely to obey | 969 | 34.0% | [32.2% - 35.7%] |
| Quite unlikely to obey | 589 | 20.7% | [19.2% - 22.1%] |
| Very unlikely to obey | 322 | 11.3% | [10.1% - 12.5%] |
| Don’t Know | 132 | 4.6% | [3.9% - 5.4%] |
| Net: Likely | 1808 | 63.4% | [61.6% - 65.2%] |
| Net: Unlikely | 911 | 32.0% | [30.2% - 33.7%] |

*Base: All adults (N=2,851)*
