## Supplementary table 5b for "Adult current and former nicotine users’ and adolescents’ views on vape-free places: findings from UK cross-sectional surveys"

**Table S5b: Perceived likelihood of people obeying rules in each location if vaping was banned: Adolescents**

| **Location** | **n** | **%** | **95% CI** |
| --- | --- | --- | --- |
| **Open air playparks** |  |  |  |
| Very likely to obey | 202 | 9.5% | [8.3% - 10.8%] |
| Quite likely to obey | 315 | 14.9% | [13.3% - 16.4%] |
| Quite unlikely to obey | 682 | 32.1% | [30.2% - 34.1%] |
| Very unlikely to obey | 750 | 35.3% | [33.3% - 37.4%] |
| Not sure | 173 | 8.1% | [7.0% - 9.3%] |
| Net: Likely | 517 | 24.4% | [22.6% - 26.2%] |
| Net: Unlikely | 1432 | 67.5% | [65.5% - 69.5%] |
| **Public transport** |  |  |  |
| Very likely to obey | 502 | 23.6% | [21.8% - 25.5%] |
| Quite likely to obey | 809 | 38.1% | [36.1% - 40.2%] |
| Quite unlikely to obey | 415 | 19.6% | [17.9% - 21.2%] |
| Very unlikely to obey | 223 | 10.5% | [9.2% - 11.8%] |
| Don’t Know | 173 | 8.2% | [7.0% - 9.3%] |
| Net: Likely | 1311 | 61.8% | [59.7% - 63.8%] |
| Net: Unlikely | 638 | 30.1% | [28.1% - 32.0%] |
| **Hospital entrances (outdoors)** |  |  |  |
| Very likely to obey | 347 | 16.3% | [14.8% - 17.9%] |
| Quite likely to obey | 555 | 26.1% | [24.3% - 28.0%] |
| Quite unlikely to obey | 626 | 29.5% | [27.5% - 31.4%] |
| Very unlikely to obey | 421 | 19.8% | [18.1% - 21.5%] |
| Don’t Know | 174 | 8.2% | [7.0% - 9.4%] |
| Net: Likely | 902 | 42.5% | [40.4% - 44.6%] |
| Net: Unlikely | 1046 | 49.3% | [47.2% - 51.4%] |
| *Base: All adolescents weighted (N=2,123).* |  |  |  |

*Due to weighting and rounding composites of ‘Likely’ and ‘Unlikely’ may not appear to add to the total of the individual items.*

**Table S5b cont’d: Perceived likelihood of people obeying rules in each location if vaping was banned: Adolescents**

| **Location** | **n** | **%** | **95% CI** |
| --- | --- | --- | --- |
| **Inside nightclubs** |  |  |  |
| Very likely to obey | 333 | 15.7% | [14.1% - 17.2%] |
| Quite likely to obey | 471 | 22.2% | [20.4% - 24.0%] |
| Quite unlikely to obey | 538 | 25.3% | [23.5% - 27.2%] |
| Very unlikely to obey | 487 | 22.9% | [21.1% - 24.7%] |
| Don’t Know | 294 | 13.9% | [12.4% - 15.3%] |
| Net: Likely | 804 | 37.9% | [35.8% - 39.9%] |
| Net: Unlikely | 1024 | 48.3% | [46.1% - 50.4%] |
| **Pubs (indoors)** |  |  |  |
| Very likely to obey | 415 | 19.6% | [17.9% - 21.3%] |
| Quite likely to obey | 664 | 31.3% | [29.3% - 33.3%] |
| Quite unlikely to obey | 493 | 23.2% | [21.4% - 25.0%] |
| Very unlikely to obey | 319 | 15.0% | [13.5% - 16.6%] |
| Don’t Know | 231 | 10.9% | [9.6% - 12.2%] |
| Net: Likely | 1080 | 50.9% | [48.7% - 53.0%] |
| Net: Unlikely | 812 | 38.2% | [36.2% - 40.3%] |
| **On school grounds (outdoors)** |  |  |  |
| Very likely to obey | 569 | 26.8% | [24.9% - 28.7%] |
| Quite likely to obey | 661 | 31.1% | [29.2% - 33.1%] |
| Quite unlikely to obey | 465 | 21.9% | [20.1% - 23.7%] |
| Very unlikely to obey | 271 | 12.8% | [11.3% - 14.2%] |
| Don’t Know | 157 | 7.4% | [6.3% - 8.5%] |
| Net: Likely | 1230 | 58.0% | [55.9% - 60.1%] |
| Net: Unlikely | 735 | 34.7% | [32.6% - 36.7%] |

*Base: All adolescents, weighted (N=2,123)	Due to weighting and rounding composites of ‘Likely’ and ‘Unlikely’ may not appear to add to the total of the individual items.*
