## Supplementary table 6a for "Adult current and former nicotine users’ and adolescents’ views on vape-free places: findings from UK cross-sectional surveys"

**Table S6a: Logistic regressions: whether think people would be unlikely to obey rules in each location: Adults**

|  |  | **a** | | | |  | **b** | | | | **c** | | | |
| --- | --- | --- | --- | --- | --- | --- | --- | --- | --- | --- | --- | --- | --- | --- |
| **Dependent variable:** |  | **Public transport** | | | |  | **On school grounds (outdoors)** | | | | **Pubs (indoors)** | | | |
|  |  | **1= Unlikely to obey ^#^ (n=681)** | | | |  | **1= Unlikely to obey ^#^ (n=911)** | | | | **1= Unlikely to obey ^#^ (n=1101)** | | | |
|  |  | **0= Likely to obey or not sure^#^ (n=2170)** | | | |  | **0= Likely to obey or not sure^#^ (n=1940)** | | | | **0= Likely to obey or not sure^#^ (n=1750)** | | | |
|  |  | **N** | **AOR*** | **95% CI** | **P** |  | **N** | **AOR*** | **95% CI** | **P** | **N** | **AOR*** | **95% CI** | **P** |
| **Gender** |  |  |  |  |  |  |  |  |  |  |  |  |  |  |
| Male |  | 1437 | Ref |  |  |  | 1437 | Ref |  |  | 1437 | Ref |  |  |
| Female |  | 1414 | 0.79 | [0.66-0.94] | 0.008 |  | 1414 | 0.93 | [0.79-1.09] | 0.361 | 1414 | 0.84 | [0.72-0.98] | 0.028 |
| **Age group** |  |  |  |  | <0.001 |  |  |  |  | 0.096 |  |  |  | <0.001 |
| 18-24 |  | 92 | Ref |  |  |  | 92 | Ref |  |  | 92 | Ref |  |  |
| 25-34 *v 18-24* |  | 548 | 0.81 | [0.50-1.31] | 0.385 |  | 548 | 1.28 | [0.77-2.11] | 0.342 | 548 | 0.73 | [0.46-1.15] | 0.177 |
| 35-44 *v 18-34* |  | 498 | 0.82 | [0.60-1.13] | 0.229 |  | 498 | 0.83 | [0.60-1.15] | 0.268 | 498 | 0.54 | [0.40-0.72] | <0.001 |
| 45-54 *v 18-44* |  | 399 | 0.88 | [0.66-1.18] | 0.393 |  | 399 | 1.17 | [0.89-1.55] | 0.260 | 399 | 0.54 | [0.41-0.70] | <0.001 |
| 55-64 *v 18-54* |  | 626 | 0.74 | [0.58-0.94] | 0.014 |  | 626 | 1.19 | [0.95-1.49] | 0.131 | 626 | 0.51 | [0.41-0.63] | <0.001 |
| 65+ *v 18-64* |  | 688 | 0.54 | [0.43-0.68] | <0.001 |  | 688 | 0.93 | [0.75-1.14] | 0.472 | 688 | 0.41 | [0.33-0.50] | <0.001 |
| **Social Grade** |  |  |  |  |  |  |  |  |  |  |  |  |  |  |
| ABC1 |  | 1979 | Ref |  |  |  | 1979 | Ref |  |  | 1979 | Ref |  |  |
| C2DE |  | 872 | 1.25 | [1.03-1.52] | 0.021 |  | 872 | 1.31 | [1.10-1.56] | 0.002 | 872 | 1.13 | [0.95-1.34] | 0.164 |
| **Country** |  |  |  |  | 0.067 |  |  |  |  | 0.152 |  |  |  | 0.057 |
| England |  | 2308 | Ref |  |  |  | 2308 | Ref |  |  | 2308 | Ref |  |  |
| Wales |  | 198 | 0.68 | [0.47-1.00] | 0.048 |  | 198 | 1.01 | [0.74-1.39] | 0.933 | 198 | 0.78 | [0.57-1.07] | 0.122 |
| Scotland |  | 258 | 0.73 | [0.53-1.01] | 0.060 |  | 258 | 1.37 | [1.05-1.79] | 0.022 | 258 | 0.75 | [0.57-1.00] | 0.047 |
| N. Ireland |  | 87 | 0.82 | [0.49-1.39] | 0.464 |  | 87 | 1.06 | [0.67-1.68] | 0.811 | 87 | 1.28 | [0.82-1.99] | 0.269 |
| **Current vaping and/or smoking status** |  |  |  |  | <0.001 |  |  |  |  | <0.001 |  |  |  | 0.212 |
| Does not currently vape or smoke |  | 482 | Ref |  |  |  | 482 | Ref |  |  | 482 | Ref |  |  |
| Dual use |  | 540 | 0.66 | [0.49-0.88] | 0.005 |  | 540 | 0.50 | [0.38-0.66] | <0.001 | 540 | 1.07 | [0.82-1.40] | 0.626 |
| Exclusively vapes |  | 1025 | 0.58 | [0.45-0.76] | <0.001 |  | 1025 | 0.50 | [0.40-0.63] | <0.001 | 1025 | 1.11 | [0.88-1.41] | 0.390 |
| Exclusively smokes |  | 804 | 0.86 | [0.66-1.11] | 0.245 |  | 804 | 0.72 | [0.57-0.91] | 0.007 | 804 | 0.90 | [0.70-1.15] | 0.407 |
| Test of model coefficients |  | χ^2^ =58.827 | | df=13 | P<0.001 |  | χ^2^ =68.877 | | df=13 | P<0.001 | χ^2^ =161.255 | | df=13 | P<0.001 |
| Naglekerke R |  | 0.031 |  |  |  |  | 0.033 |  |  |  | 0.075 |  |  |  |

*Base: All adults (N=2,851);* * adjusted for all other variables in the model, AOR, adjusted odds ratio; ref, reference category; 95% CI, 95% confidence interval. **^#^** Combines very and quite unlikely.

**Table S6a Cont’d: Logistic regressions: whether think people would be unlikely to obey rules in each location: Adults**

|  |  | **d** | | | |  | **e** | | | | **f** | | | |
| --- | --- | --- | --- | --- | --- | --- | --- | --- | --- | --- | --- | --- | --- | --- |
| **Dependent variable:** |  | **Inside nightclubs** | | | |  | **Hospital entrances (outdoors)** | | | | **Open air playparks** | | | |
|  |  | **1= Unlikely to obey ^#^ (n=1514)** | | | |  | **1= Unlikely to obey ^#^ (n=1724)** | | | | **1= Unlikely to obey ^#^ (n=2000)** | | | |
|  |  | **0= Likely to obey or not sure ^#^ (n=1337)** | | | |  | **0= Likely to obey or not sure^#^ (n=1127)** | | | | **0= Likely to obey or not sure^#^ (n=851)** | | | |
|  |  | **N** | **AOR*** | **95% CI** | **P** |  | **N** | **AOR*** | **95% CI** | **P** | **N** | **AOR*** | **95% CI** | **P** |
| **Gender** |  |  |  |  |  |  |  |  |  |  |  |  |  |  |
| Male |  | 1437 | Ref |  |  |  | 1437 | Ref |  |  | 1437 | Ref |  |  |
| Female |  | 1414 | 0.91 | [0.78-1.06] | 0.225 |  | 1414 | 1.25 | [1.07-1.45] | 0.005 | 1414 | 1.01 | [0.86-1.19] | 0.867 |
| **Age group** |  |  |  |  | <0.001 |  |  |  |  | <0.001 |  |  |  | 0.020 |
| 18-24 |  | 92 | Ref |  |  |  | 92 | Ref |  |  | 92 | Ref |  |  |
| 25-34 *v 18-24* |  | 548 | 0.30 | [0.15-0.57] | <0.001 |  | 548 | 1.17 | [0.75-1.83] | 0.491 | 548 | 0.89 | [0.55-1.43] | 0.617 |
| 35-44 *v 18-34* |  | 498 | 0.32 | [0.22-0.47] | <0.001 |  | 498 | 1.38 | [1.04-1.84] | 0.026 | 498 | 0.85 | [0.63-1.15] | 0.280 |
| 45-54 *v 18-44* |  | 399 | 0.44 | [0.32-0.59] | <0.001 |  | 399 | 1.73 | [1.33-2.26] | <0.001 | 399 | 1.21 | [0.92-1.6] | 0.174 |
| 55-64 *v 18-54* |  | 626 | 0.37 | [0.29-0.47] | <0.001 |  | 626 | 1.65 | [1.33-2.06] | <0.001 | 626 | 1.33 | [1.05-1.68] | 0.018 |
| 65+ *v 18-64* |  | 688 | 0.34 | [0.27-0.42] | <0.001 |  | 688 | 1.21 | [1.00-1.48] | 0.054 | 688 | 1.08 | [0.88-1.34] | 0.462 |
| **Social Grade** |  |  |  |  |  |  |  |  |  |  |  |  |  |  |
| ABC1 |  | 1979 | Ref |  |  |  | 1979 | Ref |  |  | 1979 | Ref |  |  |
| C2DE |  | 872 | 1.10 | [0.93-1.30] | 0.289 |  | 872 | 1.39 | [1.17-1.66] | <0.001 | 872 | 1.27 | [1.06-1.53] | 0.011 |
| **Country** |  |  |  |  | 0.298 |  |  |  |  | 0.598 |  |  |  | 0.014 |
| England |  | 2308 | Ref |  |  |  | 2308 | Ref |  |  | 2308 | Ref |  |  |
| Wales |  | 198 | 0.85 | [0.63-1.15] | 0.303 |  | 198 | 0.88 | [0.65-1.19] | 0.400 | 198 | 0.64 | [0.47-0.86] | 0.004 |
| Scotland |  | 258 | 0.81 | [0.62-1.06] | 0.125 |  | 258 | 1.05 | [0.80-1.37] | 0.741 | 258 | 1.07 | [0.80-1.44] | 0.647 |
| N. Ireland |  | 87 | 0.82 | [0.53-1.28] | 0.382 |  | 87 | 0.80 | [0.51-1.23] | 0.308 | 87 | 0.73 | [0.46-1.14] | 0.161 |
| **Current vaping and/or smoking status** |  |  |  |  | 0.066 |  |  |  |  | 0.040 |  |  |  | 0.002 |
| Does not currently vape or smoke |  | 482 | Ref |  |  |  | 482 | Ref |  |  | 482 | Ref |  |  |
| Dual use |  | 540 | 0.90 | [0.69-1.17] | 0.439 |  | 540 | 0.69 | [0.53-0.89] | 0.005 | 540 | 0.57 | [0.43-0.76] | <0.001 |
| Exclusively vapes |  | 1025 | 1.09 | [0.87-1.37] | 0.443 |  | 1025 | 0.86 | [0.68-1.08] | 0.201 | 1025 | 0.68 | [0.53-0.89] | 0.004 |
| Exclusively smokes |  | 804 | 0.85 | [0.68-1.08] | 0.188 |  | 804 | 0.85 | [0.67-1.08] | 0.192 | 804 | 0.66 | [0.51-0.87] | 0.003 |
| Test of model coefficients |  | χ^2^ =196.97 | | df=13 | P<0.001 |  | χ^2^ =90.808 | | df=13 | P<0.001 | χ^2^ =52.361 | | df=13 | P<0.001 |
| Naglekerke R |  | 0.089 |  |  |  |  | 0.042 |  |  |  | 0.026 |  |  |  |

*Base: All adults (N=2,851);* * adjusted for all other variables in the model, AOR, adjusted odds ratio; ref, reference category; 95% CI, 95% confidence interval. **^#^** Combines very and quite unlikely.
