## Supplementary table 6b for "Adult current and former nicotine users’ and adolescents’ views on vape-free places: findings from UK cross-sectional surveys"

**Table S6b: Logistic regressions: whether think people would be unlikely to obey rules in each location: Adolescents**

|  |  | **a** | | | |  | **b** | | | | **c** | | | |
| --- | --- | --- | --- | --- | --- | --- | --- | --- | --- | --- | --- | --- | --- | --- |
| **Dependent variable:** |  | **Public transport** | | | |  | **On school grounds (outdoors)** | | | | **Pubs (indoors)** | | | |
|  |  | **1= Unlikely to obey ^#^ (n=635)** | | | |  | **1= Unlikely to obey ^#^ (n=731)** | | | | **1= Unlikely to obey ^#^ (n=799)** | | | |
|  |  | **0= Likely to obey or not sure ^#^ (n=1474)** | | | |  | **0= Likely to obey or not sure ^#^ (n=1378)** | | | | **0= Likely to obey or not sure ^#^ (n=1310)** | | | |
|  |  | **N** | **AOR*** | **95% CI** | **P** |  | **N** | **AOR*** | **95% CI** | **P** | **N** | **AOR*** | **95% CI** | **P** |
| **Gender** |  |  |  |  |  |  |  |  |  |  |  |  |  |  |
| Male |  | 1136 | Ref |  |  |  | 1136 | Ref |  |  | 1136 | Ref |  |  |
| Female |  | 973 | 1.12 | [0.93-1.35] | 0.237 |  | 973 | 0.97 | [0.81-1.17] | 0.771 | 973 | 0.97 | [0.81-1.16] | 0.733 |
| **Age group** |  |  | 1.08 | [1.03-1.13] | 0.002 |  |  | 1.11 | [1.06-1.16] | <0.001 |  | 1.01 | [0.97-1.06] | 0.588 |
| **Social Grade** |  |  |  |  |  |  |  |  |  |  |  |  |  |  |
| ABC1 |  | 1545 | Ref |  |  |  | 1545 | Ref |  |  | 1545 | Ref |  |  |
| C2DE |  | 564 | 1.13 | [0.92-1.39] | 0.256 |  | 564 | 1.18 | [0.96-1.44] | 0.112 | 564 | 1.08 | [0.89-1.32] | 0.437 |
| **Country** |  |  |  |  | 0.026 |  |  |  |  | 0.098 |  |  |  | 0.086 |
| England |  | 1742 | Ref |  |  |  | 1742 | Ref |  |  | 1742 | Ref |  |  |
| Wales |  | 193 | 0.78 | [0.55-1.09] | 0.145 |  | 193 | 1.23 | [0.91-1.68] | 0.182 | 193 | 1.05 | [0.77-1.43] | 0.743 |
| Scotland |  | 103 | 1.00 | [0.64-1.56] | 0.988 |  | 103 | 1.56 | [1.03-2.36] | 0.035 | 103 | 1.31 | [0.87-1.96] | 0.198 |
| N. Ireland |  | 71 | 1.88 | [1.16-3.04] | 0.010 |  | 71 | 1.24 | [0.76-2.03] | 0.385 | 71 | 1.74 | [1.08-2.80] | 0.022 |
| **Current vaping and/or smoking status** |  |  |  |  |  |  |  |  |  |  |  |  |  |  |
| Does not currently vape |  | 1966 | Ref |  |  |  | 1966 | Ref |  |  | 1966 | Ref |  |  |
| Currently vapes |  | 143 | 1.04 | [0.72-1.50] | 0.849 |  | 143 | 1.01 | [0.71-1.45] | 0.946 | 143 | 1.28 | [0.9-1.81] | 0.170 |
| Test of model coefficients |  | χ^2^ =22.415 | | df=7 | P=0.002 |  | χ^2^ =27.559 | | df=7 | P<0.001 | χ^2^ =9.608 | | df=7 | P=0.212 |
| Naglekerke R |  | 0.015 |  |  |  |  | 0.018 |  |  |  | 0.006 |  |  |  |

*Base: All adolescents, unweighted (N=2,109), missing cases (n=14) due to missing data on social grade (n=13) and current vaping (n=1)*

* adjusted for all other variables in the model, AOR, adjusted odds ratio; ref, reference category; 95% CI, 95% confidence interval. **^#^** Combines very and quite unlikely.

**Table S6b Cont’d: Logistic regressions: whether think people would be unlikely to obey rules in each location: Adolescents**

|  |  | **d** | | | |  | **^#^ e** | | | | **f** | | | |
| --- | --- | --- | --- | --- | --- | --- | --- | --- | --- | --- | --- | --- | --- | --- |
| **Dependent variable:** |  | **Inside nightclubs** | | | |  | **Hospital entrances (outdoors)** | | | | **Open air playparks** | | | |
|  |  | **1= Unlikely to obey ^#^ (n=1011)** | | | |  | **1= Unlikely to obey ^#^ (n=1060)** | | | | **1= Unlikely to obey ^#^ (n=1432)** | | | |
|  |  | **0= Likely to obey or not sure ^#^ (n=1098)** | | | |  | **0= Likely to obey or not sure ^#^ (n=1049)** | | | | **0= Likely to obey or not sure ^#^ (n=677)** | | | |
|  |  | **N** | **AOR*** | **95% CI** | **P** |  | **N** | **AOR*** | **95% CI** | **P** | **N** | **AOR*** | **95% CI** | **P** |
| **Gender** |  |  |  |  |  |  |  |  |  |  |  |  |  |  |
| Male |  | 1136 | Ref |  |  |  | 1136 | Ref |  |  | 1136 | Ref |  |  |
| Female |  | 973 | 0.97 | [0.81-1.15] | 0.697 |  | 973 | 1.23 | [1.04-1.46] | 0.019 | 973 | 0.94 | [0.78-1.13] | 0.510 |
| **Age** |  |  | 1.04 | [0.99-1.08] | 0.110 |  |  | 1.10 | [1.05-1.15] | <0.001 |  | 1.10 | [1.05-1.15] | <0.001 |
| **Social Grade** |  |  |  |  |  |  |  |  |  |  |  |  |  |  |
| ABC1 |  | 1545 | Ref |  |  |  | 1545 | Ref |  |  | 1545 | Ref |  |  |
| C2DE |  | 564 | 0.95 | [0.78-1.15] | 0.585 |  | 564 | 1.24 | [1.02-1.51] | 0.030 | 564 | 1.18 | [0.96-1.46] | 0.122 |
| **Country** |  |  |  |  | 0.051 |  |  |  |  | 0.342 |  |  |  | 0.720 |
| England |  | 1742 | Ref |  |  |  | - | - | - | - | 1742 | Ref |  |  |
| Wales |  | 193 | 0.92 | [0.69-1.25] | 0.608 |  | - | - | - | - | 193 | 0.89 | [0.65-1.22] | 0.471 |
| Scotland |  | 103 | 1.16 | [0.78-1.73] | 0.470 |  | - | - | - | - | 103 | 1.21 | [0.78-1.88] | 0.394 |
| N. Ireland |  | 71 | 1.93 | [1.18-3.17] | 0.009 |  | - | - | - | - | 71 | 1.00 | [0.60-1.68] | 0.991 |
| **Current vaping and/or smoking status** |  |  |  |  |  |  |  |  |  |  |  |  |  |  |
| Does not currently vape |  | 1966 | Ref |  |  |  | 1966 | Ref |  |  | 1966 | Ref |  |  |
| Currently vapes |  | 143 | 1.42 | [1.01-2.01] | 0.045 |  | 143 | 0.74 | [0.52-1.04] | 0.082 | 143 | 0.67 | [0.47-0.95] | 0.026 |
| Test of model coefficients |  | χ^2^ =15.433 | | df=7 | P=0.031 |  | χ^2^ =30.069 | | df=4 | P<0.001 | χ^2^ =20.942 | | df=7 | P=0.004 |
| Naglekerke R |  | 0.010 |  |  |  |  | 0.019 |  |  |  | 0.014 |  |  |  |

*Base: All adolescents, unweighted (N=2,109), missing cases (n=14) due to missing data on social grade (n=13) and current vaping (n=1)*

* adjusted for all other variables in the model, AOR, adjusted odds ratio; ref, reference category; 95% CI, 95% confidence interval. **^#^** Combines very and quite unlikely.

**^#^** (e) Hospital entrances (outdoors): country was removed from the analysis to enable model fi
